# Effects of maternal and paternal smoking on offspring cardiometabolic risk factors in adulthood: a multi-method intergenerational Mendelian randomization study

**DOI:** 10.64898/2025.12.05.25341702

**Authors:** Grace M. Power, Tom A. Bond, Laxmi Bhatta, Bjørn Olav Åsvold, Ben Brumpton, Gibran Hemani, Deborah A. Lawlor, Geng Wang, Nicole Warrington, David M. Evans, Gunn-Helen Moen, George Davey Smith

**Affiliations:** MRC Integrative Epidemiology Unit at the University of Bristol, Bristol, UK; Population Health Sciences, Bristol Medical School, University of Bristol, Bristol, UK; Institute for Molecular Bioscience, University of Queensland, Brisbane, Australia; Frazer Institute, University of Queensland, Woolloongabba, Australia; Department of Epidemiology and Biostatistics, Imperial College London, London, UK; HUNT Center for Molecular and Clinical Epidemiology, Department of Public Health and Nursing, NTNU Norwegian University of Science and Technology, Trondheim, Norway; HUNT Research Centre, Department of Public Health and Nursing, NTNU Norwegian University of Science and Technology, Levanger, Norway; Department of Endocrinology, Clinic of Medicine, St. Olavs Hospital, Trondheim University Hospital, Trondheim 7030, Norway; Clinic of Medicine, St. Olavs Hospital, Trondheim University Hospital, Trondheim, Norway; Institute of Clinical Medicine, Faculty of Medicine, University of Oslo, Oslo, Norway; NIHR Bristol Biomedical Research Centre Bristol, University Hospitals Bristol and Weston NHS Foundation Trust, University of Bristol, Bristol, UK

## Abstract

Parental smoking has been linked to several adverse offspring cardiometabolic outcomes; however, evidence is conflicting regarding the causal and long-term nature of these associations. We investigated the effects of maternal and paternal smoking, capturing exposure before, during, and after pregnancy, on eleven offspring cardiometabolic risk factors related to body composition, blood pressure, glucose, and lipid levels in adulthood. We applied a multi-method intergenerational Mendelian randomization (MR) framework, combining two-sample MR (outcome GWAS n = up to 564,160) and one-sample MR using genetic risk score (GRS) analyses (n = up to 17,484 genotyped mother, father, offspring trios with offspring cardiometabolic risk factors) from the Norwegian HUNT cohort and British UK Biobank and ALSPAC cohorts. Smoking behaviour was instrumented using genome-wide significant variants for smoking initiation and heaviness from large genome-wide association studies (2019 and 2022), with additional analyses of the *CHRNA5* variant rs16969968.

Using the two-sample MR approach, we found that an average change in adult offspring waist-hip-ratio (WHR) per one standard deviation (SD) increase in maternal cigarettes smoked per day of 0.25 SD (95% CI: 0.13, 0.38; 2019 GWAS), 0.19 SD (95% CI: 0.08, 0.31; 2022 GWAS) and 0.27 SD (95% CI: 0.07, 0.47; *CHRNA5* rs16969968). We also found tentative evidence of comparable effects on offspring body mass index and C-reactive protein. One-sample MR analyses using *CHRNA5* rs16969968, restricted to maternal ever-smokers, provided supporting evidence for a causal effect of maternal smoking heaviness on offspring WHR. We found little evidence that maternal smoking initiation affected WHR, and little evidence that maternal smoking heaviness affected the remaining cardiometabolic risk factors; paternal smoking showed no clear effect on any outcome. Results from sensitivity analyses were consistent with these main findings.

Our finding of higher central adiposity in adult offspring of mothers with a propensity to smoke more heavily provides further incentive for prospective mothers to quit or reduce smoking heaviness before, during, and after pregnancy. Triangulating these findings using alternative methods would be valuable.

## Introduction

Cardiovascular diseases (CVDs) are the most common cause of mortality worldwide (1). The Developmental Origins of Health and Disease (DOHaD) framework postulates that the development of adult cardiovascular disease is influenced by the environment in the preconception, fertilization, embryonic, fetal, and neonatal periods (2, 3). Environmental insults in early life are hypothesised to impede development, resulting in the permanent ‘programming’ of endocrine and metabolic function (4, 5), subsequently heightening susceptibility to cardiometabolic disease in adulthood (2, 6–9).

Smoking by mothers during pregnancy is a well-established modifiable risk factor for several adverse perinatal health outcomes (10, 11). Whilst attention has largely focused on the effects of maternal smoking on conception, pregnancy, fetal and child health, there have been concerns that perinatal exposure to smoking may play an important role in increased risk of cardiometabolic disorders in offspring in their later life (11, 12). Previous conventional epidemiological analyses have shown that parental smoking is associated with increased incidence of later life obesity and related comorbidities. For example, a meta-analysis comprising 19 multivariable regression studies (n=24,201 to 308,981) found that maternal prenatal smoking was associated with higher risks of overweight, obesity, and gestational diabetes mellitus in adult offspring, but not type 2 diabetes, hypertension, waist circumference, or total cholesterol (13). In addition, an individual patient data meta-analysis which included 26 studies of mother-child-pairs (n=238,340) indicated a positive linear dose-response relationship between the number of cigarettes smoked by mothers during pregnancy and offspring overweight in childhood and adolescence for up to 15 cigarettes per day (14). An association has also been identified between paternal smoking during preadolescence and increased BMI, fat mass, and waist circumference in male offspring, with no corresponding effect observed in female offspring (15). However, a larger study conducted in the Trøndelag Health Study (HUNT), designed to reproduce these earlier findings, found little evidence that exposure to parental smoke in preadolescence was associated with offspring BMI at a mean age of 29.1 years (16). Epigenetic studies have suggested that maternal and/or paternal smoking may affect DNA methylation in the offspring, including robust evidence for persistent methylation changes following maternal smoking in pregnancy, providing a plausible mechanism by which parental smoking could increase offspring obesity risk (17, 18).

The evidence for a causal effect of parental smoking on offspring cardiometabolic risk is contradictory. A proxy gene-by-environment Mendelian randomization (MR) study found little evidence for a causal effect of maternal smoking on 12 long-term offspring outcomes, including BMI and blood pressure, although the authors acknowledged a lack of power to detect such effects (19). In addition, studies employing a sibling comparison design, which contrasts siblings with discordant exposure to maternal smoking during pregnancy (with the aim of controlling for confounders that are shared by siblings), concluded that the observed associations between maternal smoking during pregnancy and offspring obesity were more likely to be confounded by shared familial characteristics than reflect a causal intrauterine mechanism (20, 21). Earlier paternal negative control studies suggested that maternal smoking in pregnancy showed larger associations with childhood overweight and obesity than paternal smoking in mutually adjusted models, which was interpreted as being more consistent with a causal intrauterine effect (22). This study design assumes that if a causal intrauterine effect of the maternal exposure on the offspring is present, the association of the maternal exposure with the outcome should be larger than the association of the equivalent paternal exposure (23). However, evidence from an individual participant data meta-analysis of 229,158 families shows that maternal and paternal smoking have similar associations with childhood overweight, a pattern more consistent with shared familial confounding than with a specific intrauterine effect (24).

Plausible explanations for the identification of an association between maternal or paternal smoking and cardiometabolic outcomes in adult offspring include: (i) consequences from noxious chemicals found in cigarette smoke impacting the oocyte or sperm, (ii) adverse intrauterine effects from associated chemical insults, (iii) postnatal exposure to parental smoking for the offspring, or (iv) potential confounding bias. In conventional epidemiological settings, biases induced by confounding pose challenges when inferring causality. Associations identified could be due to environmental confounding or to shared genetic effects (25).

Mendelian randomization (MR) is a technique that, provided specific assumptions are met, exploits the random distribution of genetic variants from parents to offspring, independent of the influence of other traits. This reduces susceptibility to confounding factors, including confounding by undiagnosed existing disease (reverse causation) (26, 27). Recent developments in intergenerational MR methodology (28) enable indirect estimation of the effects of parental exposures on offspring adulthood outcomes by accounting for the direct fetal genotype contribution (6).

We aimed to use intergenerational MR to test the hypothesis that maternal and paternal smoking – before, during, or after pregnancy – cause a more adverse offspring cardiometabolic risk factor profile in adulthood.

## Methods

This study adheres to the Strengthening The Reporting of Observational Studies in Epidemiology Using Mendelian Randomisation (STROBE-MR) guidelines (*29, 30*) (Supplementary Material 1).

### Data sources and instruments

#### Inclusion criteria and participating cohorts

We analysed three large population-based prospective cohorts that had genome-wide SNP data available in mothers, fathers and offspring and cardiometabolic risk factors measured in adult offspring: the Trøndelag Health Study (HUNT), the UK Biobank (UKB), and the Avon Longitudinal Study of Parents and Children (ALSPAC). The selection of study participants for our main two-sample MR analysis is presented in Figure 1. Study participant selection for one-sample MR is presented in Figure S1.

**Figure 1.**
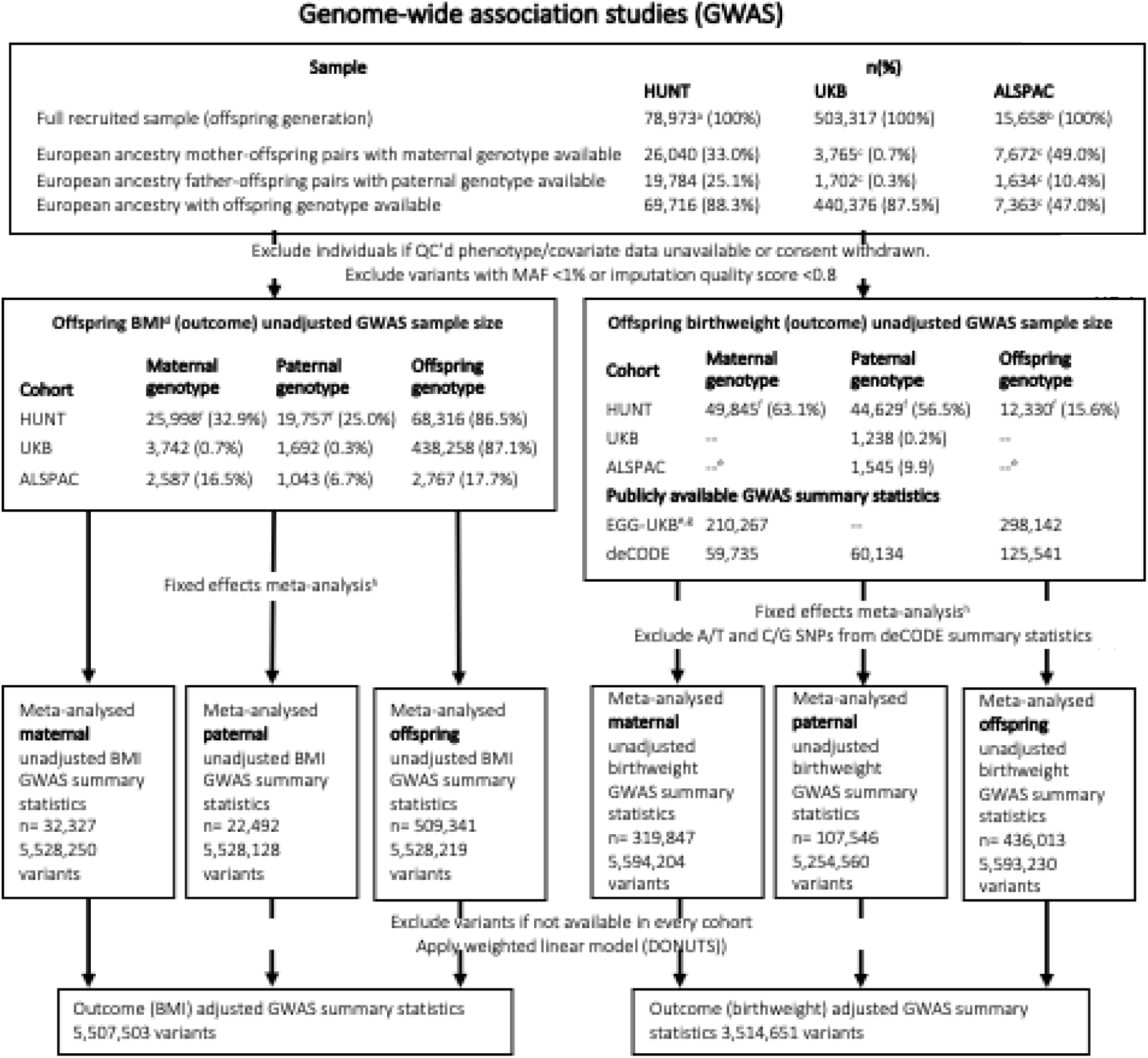
Study design overview and flowchart for instruments used in two-sample MR (GWAS) (Figure adapted from Bond, T and Bhatta, L 2025 (41, 42)). BMI: body mass index, BW: birth weight, HUNT: Trøndelag Health Study, UKB: UK Biobank, ALSPAC: Avon Longitudinal Study of Parents and Children, EGG: Early Growth Genetics Consortium, GIANT: Genetic Investigation of Anthropometric Traits Consortium, GWAS: genome-wide association study, QC: quality control, MAF: minor allele frequency. BW: birth weight, HUNT: Trøndelag Health Study, UKB: UK Biobank, ALSPAC: Avon Longitudinal Study of Parents and Children, EGG: Early Growth Genetics Consortium, GIANT: Genetic Investigation of Anthropometric Traits Consortium, GWAS: genome wide association study, QC: quality control, MAF: minor allele frequency. a: total number of participants in the HUNT2 and HUNT3 study waves, b: total number of fetuses included in ALSPAC (including participants recruited after 7 years of age), c: only unrelated individuals within the parent and offspring generations were included in the designated samples, d: sample sizes were similar for the other adult outcomes, e: ALSPAC data were included in EGG, f: in HUNT, the parent-offspring samples included siblings in the offspring generation, and for the birth weight analyses parent-offspring pairs were identified via a birth registry instead of via genetic data, resulting in larger samples for parental GWAS and a smaller sample for offspring GWAS compared with adult outcomes, g: we used publicly available summary statistics from the meta-analysis of EGG, UKB and deCODE carried out by Juliusdottir *et al*. (30), h: prior to meta-analysis, A/T and C/G SNPs were removed when comparison of their allele frequency to the HRC or 1000 Genome Project reference panel suggested harmonisation errors.

##### HUNT

HUNT is a cohort study of the adult population in Trøndelag County, Norway, and comprises ∼230,000 individuals aged ≥20 years (31, 32). Individuals were recruited in four surveys from 1984–2019, of which the HUNT2 and HUNT3 surveys were included in the present study. Invitee participation rates were 69% and 54%, respectively. Data on perinatal outcomes are available via linkage to the Medical Birth Registry of Norway (MBRN) (33) for individuals born after 1967. A total of 69,716 European ancestry offspring were available in HUNT, with genotype data available, of which 26,040 had maternal genotype data available and 19,784 had psaternal genotype data available (6, 34, 35). Parent-offspring pairs and trios were identified using kinship analysis implemented in the KING software package (36).

##### UKB

UKB is a cohort of 503,317 adult volunteers, recruited from across the UK aged 40–69 years between 2006 and 2010 (37). Parent-offspring pairs and trios were identified using kinship analysis implemented in the KING software package (36).

##### ALSPAC

ALSPAC is a birth cohort which enrolled 14,541 pregnant women living in Avon, England with expected dates of delivery between April 1, 1991, and December 31, 1992 (the estimated recruitment rate of those eligible was 80%) (38, 39). Further enrolments after 1998 resulted in a baseline sample of 14,901 children alive at one year of age. We used data collected during pregnancy/birth and the 24-year follow up. Some study data were collected and managed using Research Electronic Data Capture (REDCap) electronic data capture tools hosted at the University of Bristol (40). Please note that the study website contains details of all the data that is available through a fully searchable data dictionary and variable search tool: http://www.bristol.ac.uk/alspac/researchers/our-data/. Parent-offspring pairs and trios were identified by matching on study ID variables.

Informed consent was obtained from all participants and ethical approval was obtained from the Regional Committee for Medical and Health Research Ethics, Central Norway (REK Central application number 2018/2488) (HUNT), the North West Multi-centre Research Ethics Committee (MREC) (ref 11/NW/0382) (UKB), and the ALSPAC Ethics and Law Committee and the Local Research Ethics Committees (ALSPAC).

#### Offspring cardiometabolic risk factors and birthweight

Standard protocols were used to assess the eleven offspring cardiometabolic outcomes in adulthood (body mass index (BMI), waist-hip-ratio (WHR), systolic (SBP) and diastolic (DBP) blood pressure, blood glucose, glycated haemoglobin (HbA1c), total cholesterol, high density lipoprotein cholesterol (HDL-C), low density lipoprotein cholesterol (LDL-C), triglycerides and high sensitivity C-reactive protein (CRP)) (41, 42). For one-sample MR analyses, CRP was only available in ALSPAC and UKB and HbA1c was only available in UKB. Blood pressure and lipid measurements were corrected for antihypertensive and lipid lowering medication use as described previously (41, 42). Briefly, blood pressure values for participants in HUNT and UKB who reported taking antihypertensive medication were corrected by adding 15 mmHg and 10 mmHg to the raw SBP and DBP values respectively. Lipid values were corrected for participants reporting taking lipid lowering medication in UKB by dividing the raw LDL cholesterol values by 0.7 and the raw total cholesterol and triglyceride values by 0.8 (41, 42). Biochemical values were measured in fasting blood samples in ALSPAC and non-fasting in HUNT and UKB. Offspring birthweight was employed as a positive control outcome in this analysis. A priori, we expected to see a negative causal effect of smoking during pregnancy on birthweight (43–45). Birthweight was extracted from the Medical Birth Registry of Norway (MBRN) in HUNT, measured by trained research assistants or abstracted from the birth record/notification in ALSPAC, and retrospectively self-reported at recruitment in UKB. CRP was strongly right skewed, therefore was natural log transformed prior to GWAS.

#### Genotyping, quality control and imputation in each cohort

Genotyping, quality control and imputation in each cohort are described in detail in the Supplementary Material 2 and elsewhere (34, 46–48). To summarise, participants were genotyped using genome-wide arrays, followed by imputation of genome-wide variants. HUNT participants were imputed using a customised Haplotype Reference Consortium (HRC) panel including 2,201 Norwegian whole genome sequences. ALSPAC mothers and offspring were imputed to the HRC panel (34, 47), and ALSPAC fathers to the 1000 Genomes phase 1 panel (48). UKB participants were imputed to a merged HRC/UK10K panel (46). Imputed variants with minor allele frequency (MAF) >1% and imputation accuracy score >0.8 were used for genome wide association study (GWAS) analyses.

#### Parental smoking exposures

We explored effects of maternal and paternal smoking initiation (by using genetic variants associated with self-reported ever versus never smokers), intensity of smoking (by using genetic variants associated with self-reported number of cigarettes per day the rs16969968 genetic variant in the *CHRNA5* gene shown to act primarily via smoking heaviness (49, 50)). Thus, the measures we used aim to capture whether:

(i) current smoking compared to never smoking causes an effect on the outcomes of interest, and
(ii) the intensity of smoking on the outcomes of interest suggests a dose-response relationship.

This study utilised external genome-wide association studies (GWASs) carried out by the GWAS and Sequencing Consortium of Alcohol and Nicotine use (GSCAN) in 2019 and 2022 (Table S1) (51, 52). We use both 2019 and 2022 GWAS. Larger GWAS typically identify variants with smaller effect sizes, that may be more prone to pleiotropy if the primary phenotype is imprecisely defined. Consequently, the smaller 2019 GWAS may yield estimates that are less affected by pleiotropic bias. In addition, *CHRNA5* (which is a region containing the strongest signal for smoking heaviness) (49, 50) was investigated using rs16969968 as a single instrumental variable, sourced from the above 2022 GWAS of cigarettes per day (52). In addition, there is weak association of this variant with never and ever smoking, thus allowing stratification by these groups without introducing substantial collider bias (53). The GWAS selected were conducted on participants of European ancestry to mitigate bias by population stratification.

### Statistical analysis

When genetic variants are used as instrumental variables in MR, the assumptions of instrumental variables must be met, i.e., the genetic variants must (i) be strongly associated with the exposure of interest and relevant to the population studied, here pregnant women (“relevance”), (ii) not share common causes with the outcome (“independence”), and (iii) not affect the outcome other than through the exposure (“exclusion-restriction”) (54).

To block the path from parental genotype through offspring genotype to the offspring outcome in an intergenerational context (which would be a violation of the “exclusion restriction” assumption), MR is robust if (i) summary outcome associations are extracted from adjusted GWAS (parental genotype adjusted for offspring genotype and if relevant, the other parent’s genotype) in a two-sample setting, or (ii) the association between maternal or paternal unweighted GRS and offspring outcomes is adjusted for offspring genotype (and if relevant, other parent GRS), in a one-sample setting (6, 28, 55, 56). There are other valid ways to conduct intergenerational one-sample MR (e.g. maternal non-transmitted alleles) (57), which will not be used in this current study.

#### Summary level two-sample Mendelian randomisation

For the main analysis, we conducted two-sample intergenerational MR using previously derived GWAS summary statistics for each cardiometabolic risk factor (combined n for maternal, paternal and offspring GWAS = up to 564,160), which employed maternal and paternal genotype, and adjusted for the offspring’s genotype (41, 42). These were generated using a weighted linear modelling (WLM) approach, implemented in the DONUTS R package, accounting for the correlation between the parental genotypes (58). The WLM estimated the adjusted genetic effects (i.e. the mutually adjusted coefficients for maternal, paternal and offspring genotype, fitted jointly in the same model) as linear combinations of the unadjusted genetic effects (i.e. the coefficients for maternal, paternal and offspring genotype fitted in separate, partially overlapping samples) (55, 59, 60). We use the shorthand “unadjusted” to refer specifically to models that are unadjusted for parental/offspring genotype, as opposed to the other covariates, inclufing offspring age, sex, and genetic principal components, which were included in all models. We found that MR estimates changed very little when using genetic effects from a “duos” WLM (that only adjusted for offspring genotype) compared with estimates from a “trios” WLM (that adjusted for offspring genotype and the other parent’s genotype). Since the duos WLM yields smaller standard errors, we present the duos results as our primary analysis (trios results are presented in the supplementary information). GWASs were carried out on all outcomes using mother, father, and offspring genotypes. We assume that imputation quality is the same across maternal, paternal and offspring GWAS. Methods are described in detail elsewhere (41, 42). In brief, offspring outcomes were regressed separately on maternal, paternal or offspring imputed genotype dosages, adjusting for offspring sex, age at outcome measurement (including linear, quadratic and sex interaction effects, for all outcomes except birthweight), technical covariates and the top 20 genetic principal components (calculated from maternal, paternal or offspring genotypes respectively) to adjust for population stratification. A linear mixed model using the fastGWA method (61) was used to account for cryptic relatedness. Parent-offspring pairs of European ancestry, for whom parental genotype and offspring phenotype data were available, were used. These analyses were conducted with HUNT, UKB, and ALSPAC cohorts independently and then meta-analysed using a fixed effects model implemented in METAL version 2011-03-25 (62). Standard quality control (QC) procedures were carried out including calculating the Linkage Disequilibrium Score Regression (LDSC) intercept and attenuation ratio.

To estimate the causal maternal (Figure 2A) and paternal (Figure 2B) effects of smoking initiation and intensity on offspring outcomes we first used two-sample intergenerational MR, with the SNP-exposure estimates from the GSCAN GWAS and the SNP-outcome estimates from the adjusted maternal/paternal/offspring GWAS. We computed the inverse variance weighted (IVW) estimate which takes a weighted mean of the variant-specific causal estimates using inverse variance weights (63). In IVW analyses, the regression slope (ratio) is forced through a zero intercept. This assumes balanced horizontal pleiotropy. We additionally ran sensitivity analyses using MR-Egger, weighted median, and weighted mode methods (64–66). MR-Egger evaluates the potential presence of pleiotropic effects by allowing for a non-zero intercept in the regression of the variant-outcome estimates on variant-exposure estimates (64). It provides an unbiased estimate of the causal effect even in the presence of horizontal pleiotropy assuming that the INstrument Strength Independent of Direct Effect (InSIDE) assumption holds (67). The weighted median method provides an alternative estimate of the causal effect by considering the weighted median rather than the weighted mean of the instrumental variable estimates (65). We checked for evidence of horizontal pleiotropy via Cochran’s Q test for heterogeneity, and by testing whether the MR Egger intercept differed from zero. There is evidence that genetic variants associated with smoking also influence body composition, suggesting potential shared genetic pathways between smoking behaviours and adiposity (68, 69). Smoking-associated genetic variants are also correlated with socioeconomic position, commonly indexed by educational attainment, indicating that these variants may capture both behavioural and socioeconomic influences (52, 70, 71). To account for these overlapping genetic relationships, we applied multivariable MR (MVMR) in our two-sample analyses to estimate the effect of smoking initiation and smoking intensity on WHR, conditional on educational attainment and BMI. By modelling these exposures jointly, MVMR provides smoking-WHR estimates that account for these shared genetic pathways, under the multivariable IV assumptions (72, 73). Instrument strength in the multivariable MVMR models was evaluated using conditional F-statistics, which account for the inclusion of multiple, potentially correlated exposures. These analyses were performed using the *TwoSampleMR* R package (58).

**Figure 2.**
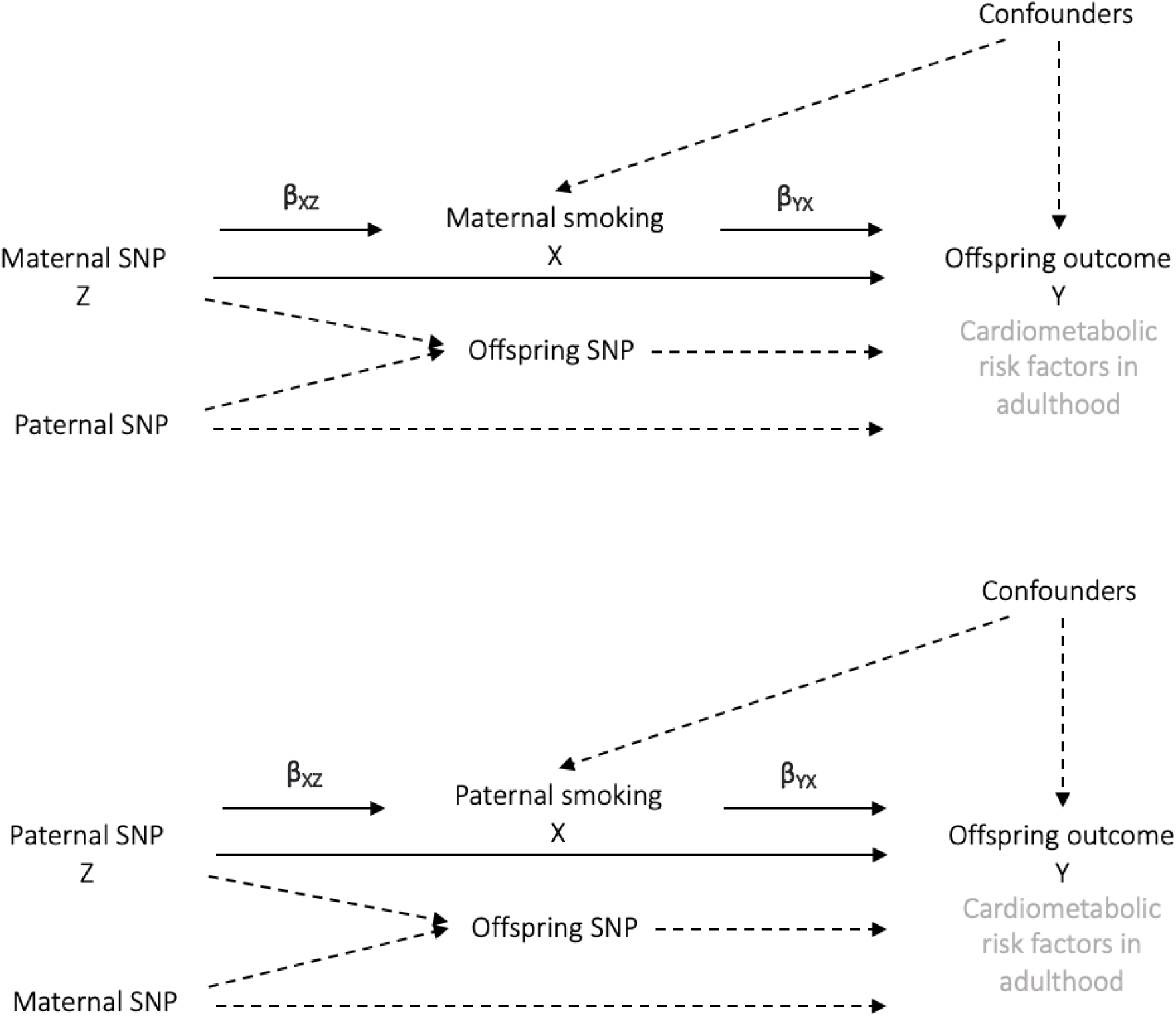
Directed acyclic graphs indicating two scenarios to explain the causal effect of parental smoking on offspring cardiometabolic risk factors in adulthood. A. Intergenerational MR estimating maternal smoking on offspring cardiometabolic risk factors in adulthood. The offspring and paternal SNP are adjusted for and thus dotted paths are blocked B. Intergenerational MR estimating paternal smoking on offspring cardiometabolic risk factors in adulthood. The offspring and maternal SNP are adjusted for and thus dotted paths are blocked.

Results subsequently underwent adjustment for multiple testing using Bonferroni correction for 12 outcomes (corrected significance level p=0.004) to protect against Type 1 error (74). Details of the input parameters are presented in Table S2.

#### Individual-level one-sample MR using GRS analysis

In one-sample MR, we explored effects of maternal and paternal intensity of smoking using instruments identified in the GWAS described previously (51, 52). These one-sample analyses represent reduced form associations, where offspring outcomes are regressed directly on parental genetic scores without scaling by the genetic association with smoking exposure. Accordingly, offspring outcomes were regressed separately on maternal, paternal, or offspring genetic instruments, assuming an additive genetic model. Results therefore reflect the overall genetic-outcome association, not the causal effect per unit change in the smoking exposure. Unweighted genetic scores were constructed by summing smoking-raising alleles (Table S3). We additionally ran these analyses using just the variant rs16969968 in *CHRNA5*. We examined the association between the parental GRS of interest and offspring outcomes, adjusting for the offspring’s genetic score (and the other parent’s GRS) calculated from the same smoking heaviness-associated SNPs. These analyses were conducted in up to 17,484 mother-father-offspring trios from HUNT, UKB and ALSPAC.

We constructed an unweighted GRS since regression coefficients obtained in the smoking GWAS’s might not accurately reflect effect sizes during pregnancy (75, 76). We fitted a linear mixed model implemented in the GCTA software package (77) using a genetic relatedness matrix (GRM) constructed from the offspring genotypes to account for relatedness and population stratification. Known smoking-associated SNPs and any SNPs within ±1 Mb of these loci (to account for linkage disequilibrium) were excluded from GRM construction (6). SNPs included in GRSs were selected from GWAS meta-analyses as genome-wide significant (P < 5 × 10⁻⁸) and conditionally independent, based on LD clumping or fine-mapping procedures described by the original studies (51, 52). All analyses were adjusted for offspring age at measurement and sex. The GRM mitigated the need to adjust for PCs. Parent-offspring pairs of European ancestry, for whom parental genotype and offspring phenotype data were available, were used. We implemented a complete-case approach, excluding individuals with missing values from the analysis. These analyses were conducted in HUNT, UKB and ALSPAC and then meta-analysed. A fixed-effects meta-analysis of regression coefficients from the three cohorts was conducted using Stata version 16 (78).

#### Stratified analysis

If our genetic instruments for smoking heaviness (including rs16969968 in the *CHRNA5* gene) are valid, we would expect these variants to only indicate a causal effect when the mother/father is a smoker. If a causal effect is detected in parental never-smokers, this implies that the *CHRNA5* gene is influencing the offspring outcome via mechanisms other than smoking heaviness (i.e. horizontal pleiotropy). We therefore repeated the *CHRNA5* gene MR analyses, stratifying the sample to parental ever-smokers and never-smokers (49). For two sample analyses we only had a sufficiently large sample to run stratified analyses for birthweight (because the WLM relies on LD score regression (79), which has stringent sample size requirements), whereas we ran stratified one-sample analysis for adult outcomes too.

## Results

### Summary level two-sample Mendelian randomisation

The GWAS meta-analyses used for the SNP-outcome association in the two-sample MR included between 14,481 (for HbA1c with paternal genotype) and 511,253 (for DBP with offspring genotype) participants (Table S4) (80, 81).

We found strong evidence of an average change in adult offspring WHR per one standard deviation (SD) increase in maternal cigarettes smoked per day of 0.25 SD (95% CI: 0.13, 0.38) using GWAS carried out in 2019, 0.19 SD (95% CI: 0.08, 0.31) using the GWAS carried out in 2022, and 0.27 SD (95% CI: 0.07, 0.47) using *CHRNA5* rs16969968 (Figure 3A, Table S5). The estimates presented here all derive from unstratified analyses; stratified analyses are presented later. There was weaker evidence that increased maternal smoking heaviness had a causal effect on higher BMI (average change in SD of BMI in adult offspring per maternal cigarettes smoked per day of 0.23 SD (95% CI: 0.07, 0.39) using GWAS carried out in 2019, 0.14 SD (95% CI: 0.00, 0.29) using GWAS carried out in 2022 and 0.11 SD (95% CI: −0.13, 0.35) using *CHRNA5* rs16969968) and CRP (average change in SD of CRP in adult offspring 1SD greater maternal cigarettes smoked per day of 0.24 SD (95% CI: 0.08, 0.40) using GWAS carried out in 2019, 0.20 SD (95% CI: 0.04, 0.36) using GWAS carried out in 2022, and 0.25 SD (95% CI: 0.01, 0.49) using *CHRNA5* rs16969968). Corresponding paternal effect estimates are presented in Table S6. In the analyses using the trios WLM for the outcome GWAS (in which the parental genetic effects were adjusted for the other parent’s genotype as well as the offspring’s genotype), results were similar, although the confidence intervals were wider due to the lower statistical power of the trios WLM versus the duos WLM (Figure S2).

**Figure 3.**
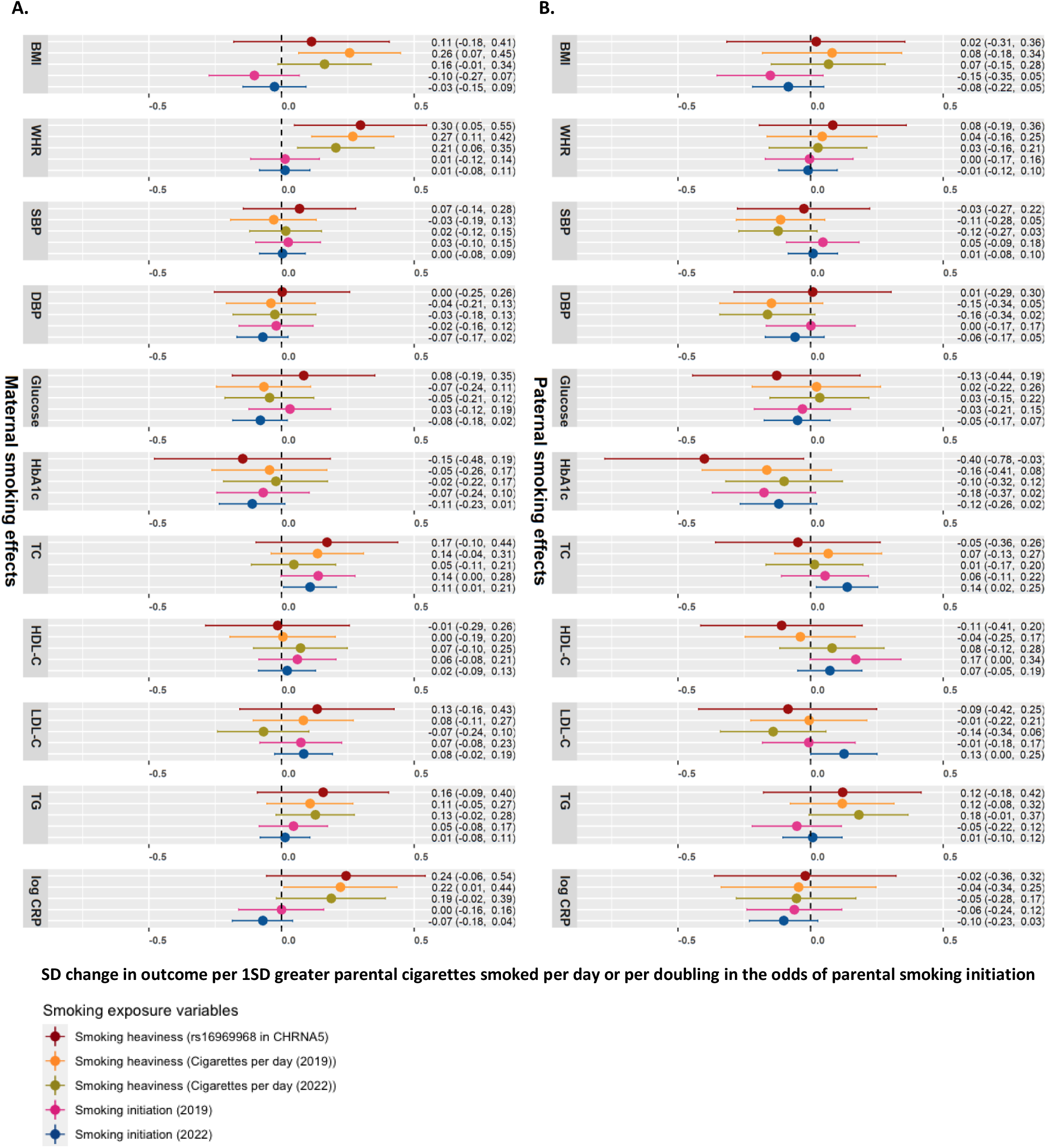
Two-sample MR for maternal and paternal smoking heaviness and initiation onto 11 long term cardiometabolic outcomes in offspring. Parental genotype was adjusted for offspring genotype (duos WLM). A. Maternal smoking on offspring cardiometabolic risk factors in adulthood. B. Paternal smoking on offspring cardiometabolic risk factors in adulthood. MR estimates for the effect of smoking heaviness can be interpreted as the average change in the outcome, in standard deviation (SD) units, per one SD greater parental cigarettes smoked per day. MR estimates for the effect of smoking initiation can be interpreted as the average change in the outcome, in SD units, per doubling in the odds of smoking initiation.

Point estimates from the two-sample weighted median, weighted mode and MR-Egger methods were similar to IVW estimates and evidence remained robust for the maternal effect for smoking heaviness on offspring WHR in later life (Table S5). In addition, Cochran’s Q test provided little evidence for between-SNP heterogeneity, and there was not strong evidence that the MR Egger intercept differed from zero (Table S7). This suggests little evidence that horizontal pleiotropy is driving these results. After correcting for multiple testing using a Bonferroni test, evidence of an effect remained for maternal smoking heaviness on increased WHR and BMI only (using p=0.004). Findings for adult offspring outcomes using the unadjusted GWAS sensitivity analyses are presented in Table S8 for maternal exposures and Table S9 for paternal exposures. MVMR analyses indicated that there remained strong evidence for the effect of maternal smoking heaviness on WHR in offspring in later life after accounting for BMI as well as educational attainment (Table S10). There was little evidence of an effect of either educational attainment or BMI on WHR in offspring after adjustment for any maternal smoking indicator. Comparison of the conditional F-statistics showed that the instruments for BMI and educational attainment were substantially stronger than those for the smoking exposures (Table S10). The strength of the BMI and educational attainment instruments was largely preserved when conditioned on smoking, indicating that these estimates were not affected by SNP non-overlap. In contrast, conditioning led to considerable weakening of the smoking instruments, consistent with the shared genetic architecture between smoking behaviour and BMI. Accordingly, estimates for smoking heaviness should be interpreted with some caution; however, the persistence of an association with offspring WHR after adjustment for BMI suggests that this effect is not solely attributable to shared genetic pathways and may in fact be attenuated by weak-instrument bias.

### Individual-level one-sample MR using GRS analysis

The individual-level one-sample GRS analysis comprised between 16,942 and 17,484 mother-father-offspring trios (Table 1 and Table S11). These one-sample analyses represent reduced form associations between parental genetic scores and offspring outcomes. One-sample MR in the total population indicated that higher genetically predicted maternal smoking heaviness (using rs16969968 in the *CHRNA5* SNP) had a positive effect on WHR, TC, and LDL-cholesterol in offspring in later life (Figure 4A, Table S12). There was no evidence of paternal smoking effects (Figure 4B, Table S13). Cohort-specific results are presented in Tables S14-S16. Effect estimates obtained using the unweighted smoking GRS were smaller than those from rs16969968 in *CHRNA5*, consistent with differences in instrument scale: a single *CHRNA5* allele has a larger established effect on smoking intensity than a one-allele change in the unweighted multi-locus score. Consequently, the unweighted GRS yields more diluted reduced-form associations.

**Figure 4.**
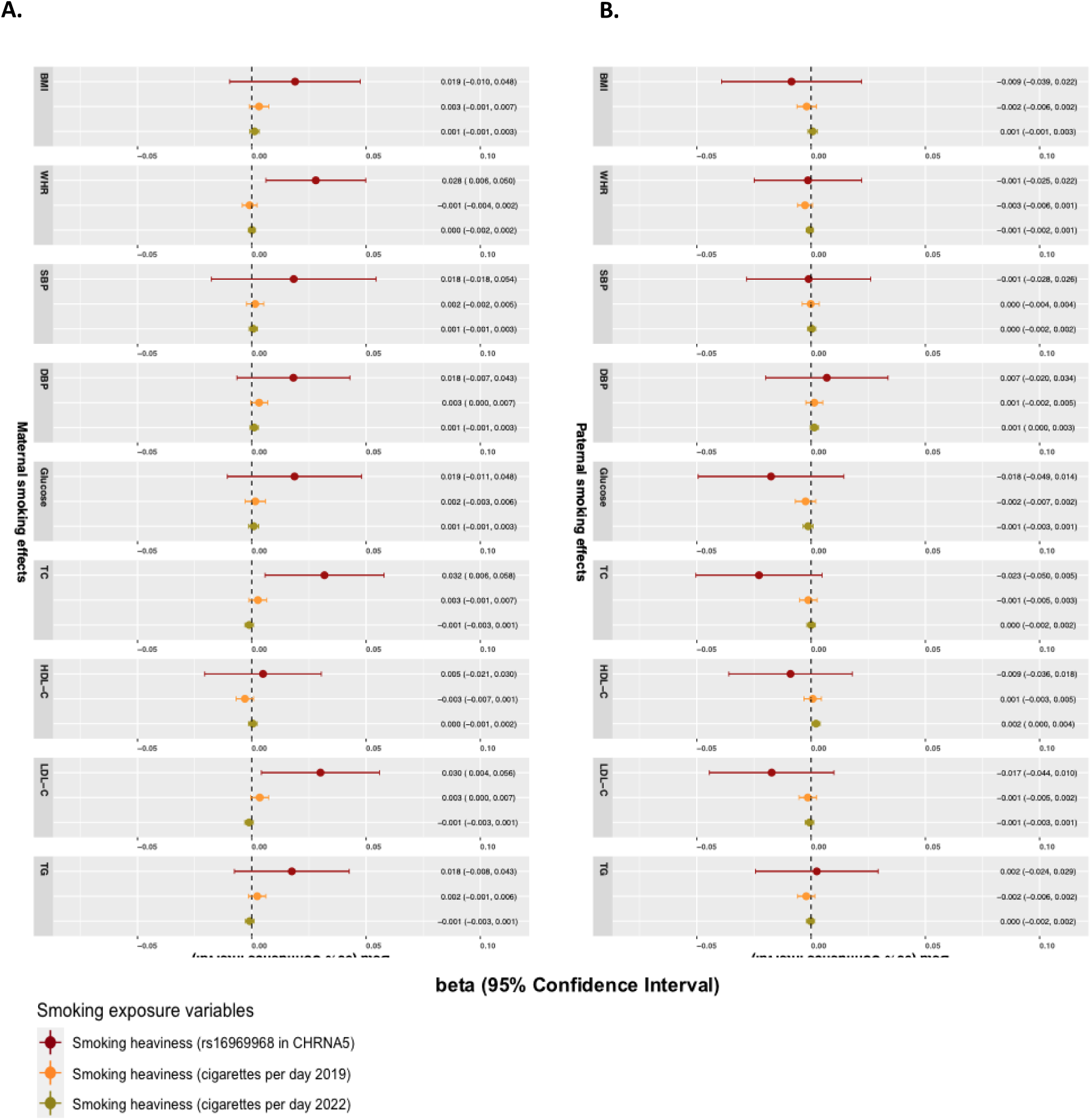
One-sample MR for smoking heaviness onto 9 long term cardiometabolic outcomes in offspring. A. Maternal smoking on offspring cardiometabolic risk factors in adulthood. B. Paternal smoking on offspring cardiometabolic risk factors in adulthood. Estimates represent the average change in offspring cardiometabolic outcomes per additional effect allele in the unweighted smoking GRS and per copy of the CHRNA5 risk allele, expressed in standard deviation (SD) units of each outcome. These estimates reflect associations between the parental genetic instruments and offspring outcomes, rather than causal effects per unit change in smoking heaviness.

**Table 1.**
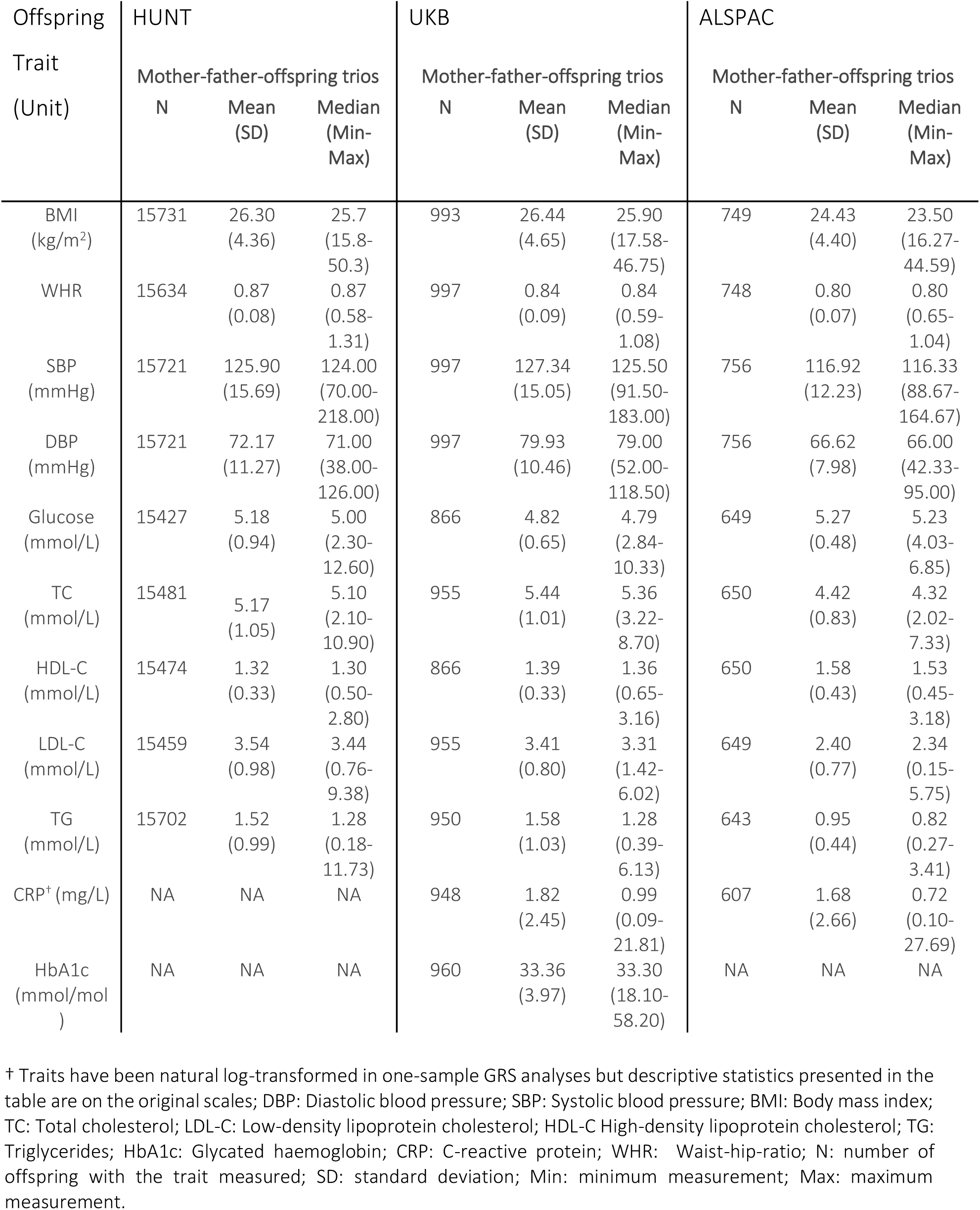
Descriptive statistics of the offspring’s cardiometabolic traits for mother-father-offspring trios in HUNT, UKB and ALSPAC.

| Offspring<br>Trait<br>(Unit) | HUNT |  |  | UKB |  |  | ALSPAC |  |  |
| --- | --- | --- | --- | --- | --- | --- | --- | --- | --- |
|  | Mother-father-offspring trios |  |  | Mother-father-offspring trios |  |  | Mother-father-offspring trios |  |  |
|  | N | Mean<br>(SD) | Median<br>(Min-Max) | N | Mean<br>(SD) | Median<br>(Min-Max) | N | Mean<br>(SD) | Median<br>(Min-Max) |
| BMI<br>(kg/m<sup>2</sup>) | 15731 | 26.30<br>(4.36) | 25.7<br>(15.8-50.3) | 993 | 26.44<br>(4.65) | 25.90<br>(17.58-46.75) | 749 | 24.43<br>(4.40) | 23.50<br>(16.27-44.59) |
| WHR | 15634 | 0.87<br>(0.08) | 0.87<br>(0.58-1.31) | 997 | 0.84<br>(0.09) | 0.84<br>(0.59-1.08) | 748 | 0.80<br>(0.07) | 0.80<br>(0.65-1.04) |
| SBP<br>(mmHg) | 15721 | 125.90<br>(15.69) | 124.00<br>(70.00-218.00) | 997 | 127.34<br>(15.05) | 125.50<br>(91.50-183.00) | 756 | 116.92<br>(12.23) | 116.33<br>(88.67-164.67) |
| DBP<br>(mmHg) | 15721 | 72.17<br>(11.27) | 71.00<br>(38.00-126.00) | 997 | 79.93<br>(10.46) | 79.00<br>(52.00-118.50) | 756 | 66.62<br>(7.98) | 66.00<br>(42.33-95.00) |
| Glucose<br>(mmol/L) | 15427 | 5.18<br>(0.94) | 5.00<br>(2.30-12.60) | 866 | 4.82<br>(0.65) | 4.79<br>(2.84-10.33) | 649 | 5.27<br>(0.48) | 5.23<br>(4.03-6.85) |
| TC<br>(mmol/L) | 15481 | 5.17<br>(1.05) | 5.10<br>(2.10-10.90) | 955 | 5.44<br>(1.01) | 5.36<br>(3.22-8.70) | 650 | 4.42<br>(0.83) | 4.32<br>(2.02-7.33) |
| HDL-C<br>(mmol/L) | 15474 | 1.32<br>(0.33) | 1.30<br>(0.50-2.80) | 866 | 1.39<br>(0.33) | 1.36<br>(0.65-3.16) | 650 | 1.58<br>(0.43) | 1.53<br>(0.45-3.18) |
| LDL-C<br>(mmol/L) | 15459 | 3.54<br>(0.98) | 3.44<br>(0.76-9.38) | 955 | 3.41<br>(0.80) | 3.31<br>(1.42-6.02) | 649 | 2.40<br>(0.77) | 2.34<br>(0.15-5.75) |
| TG<br>(mmol/L) | 15702 | 1.52<br>(0.99) | 1.28<br>(0.18-11.73) | 950 | 1.58<br>(1.03) | 1.28<br>(0.39-6.13) | 643 | 0.95<br>(0.44) | 0.82<br>(0.27-3.41) |
| CRP<sup>†</sup> (mg/L) | NA | NA | NA | 948 | 1.82<br>(2.45) | 0.99<br>(0.09-21.81) | 607 | 1.68<br>(2.66) | 0.72<br>(0.10-27.69) |
| HbA1c<br>(mmol/mol<br>) | NA | NA | NA | 960 | 33.36<br>(3.97) | 33.30<br>(18.10-58.20) | NA | NA | NA |

† Traits have been natural log-transformed in one-sample GRS analyses but descriptive statistics presented in the table are on the original scales; DBP: Diastolic blood pressure; SBP: Systolic blood pressure; BMI: Body mass index; TC: Total cholesterol; LDL-C: Low-density lipoprotein cholesterol; HDL-C High-density lipoprotein cholesterol; TG: Triglycerides; HbA1c: Glycated haemoglobin; CRP: C-reactive protein; WHR: Waist-hip-ratio; N: number of offspring with the trait measured; SD: standard deviation; Min: minimum measurement; Max: maximum measurement.

### Stratified analysis

For GWAS of birthweight stratified by parental smoking status, analysis comprised between 10,719 and 136,933 participants in maternal ever smokers, 8,094 and 104,002 participants in maternal never smokers, 9,425 and 27,726 participants in paternal ever smokers and 5,367 and 18,718 participants in paternal never smokers (Table S17). In analyses employing the unadjusted GWAS (i.e. using genetic effects that were not adjusted for the offspring’s or other parent’s genotype), in the unstratified sample (including parental never and ever smokers) we found suggestive evidence that maternal and paternal smoking initiation caused lower birthweight (Figure 5, Table S18). We also found some weak evidence that greater maternal smoking heaviness caused lower offspring birth weight. However, in the maternal ever smoker group these effects were noticeably stronger. Interestingly, in the maternal never smoker group the effects of maternal smoking heaviness flipped sign and became weakly positive. Taken together, these observations support our use of birth weight as a positive control outcome. In the analyses using the adjusted GWAS, the direction of effect was often consistent with analyses using the unadjusted GWAS, but the adjusted GWAS had lower statistical power, resulting in weaker evidence (Figure S3, Figure S4). There was little evidence that paternal smoking heaviness influenced offspring birth weight (Figure 5, Figure S3; Table S18).

**Figure 5.**
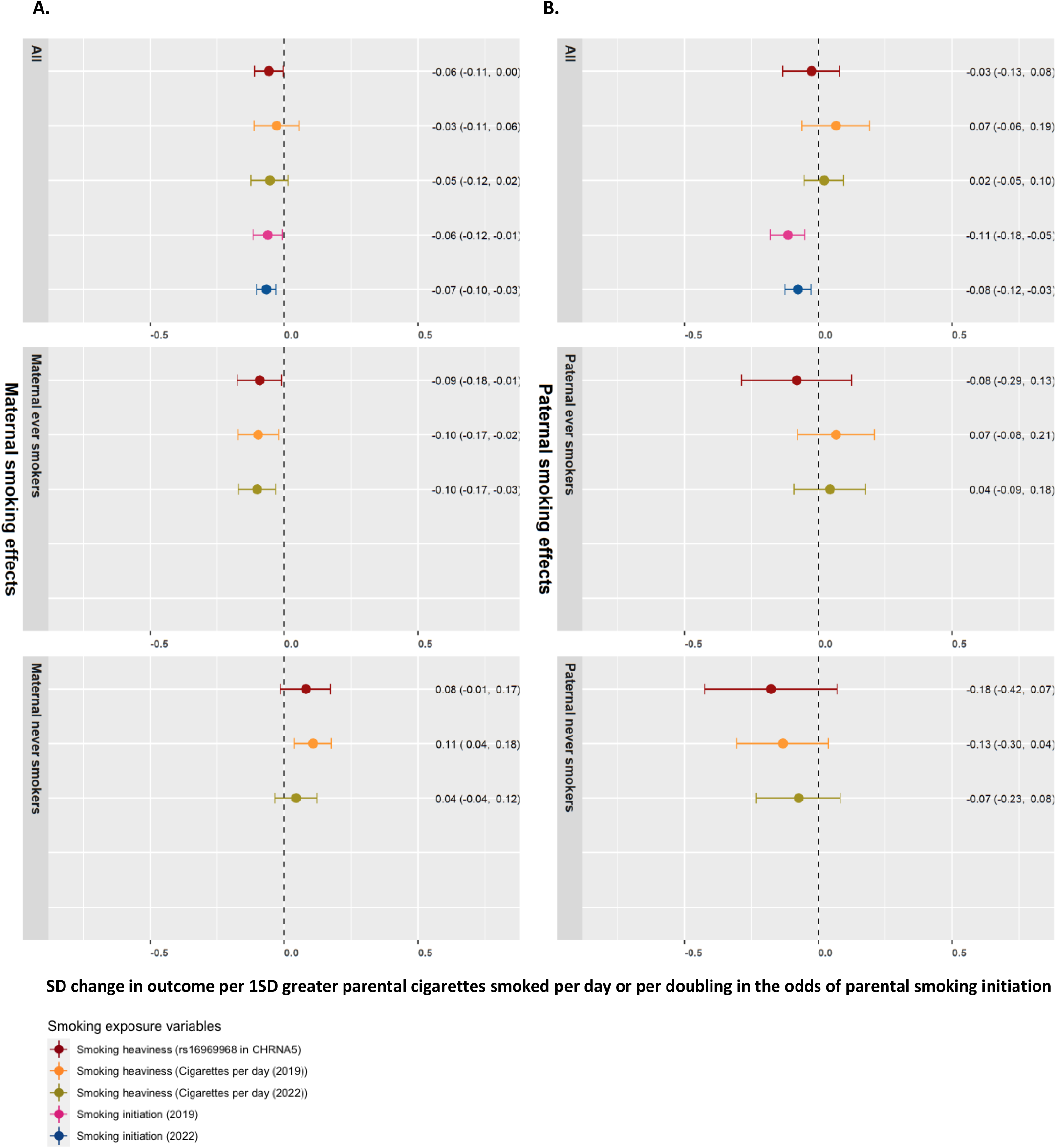
Two-sample MR for maternal and paternal smoking heaviness and initiation onto birthweight in offspring, stratified by parental smoking status, using unadjusted (marginal) genetic effects. A. Maternal smoking exposures on birthweight. B. Paternal smoking exposures on birthweight. MR estimates for the effect of smoking heaviness can be interpreted as the average change in the outcome, in standard deviation (SD) units, per one SD greater parental cigarettes smoked per day. MR estimates for the effect of smoking initiation can be interpreted as the average change in the outcome, in SD units, per doubling in the odds of smoking initiation.

The one-sample MR stratified smoking samples comprised between 8,814 and 9,065 participants in maternal ever smokers, 7,357 and 7,624 in maternal never smokers, 12,004 and 12,339 in paternal ever smokers and 4,445 and 4,623 in paternal never smokers, with numbers differing across outcomes (Table S11). Whilst the sample size was smaller and thus power was low, the magnitude of effect became slightly stronger in ever smokers for the effects of maternal smoking heaviness instrumented by rs16969968 in *CHRNA5* on WHR and maternal smoking heaviness instrumented by cigarettes per day on BMI (Figure 6A, Table S12). There was additionally some evidence of an effect of maternal smoking heaviness instrumented by rs16969968 in *CHRNA5* on DBP and SBP in ever smokers. There was weak evidence of an effect of maternal smoking heaviness instrumented by rs16969968 in *CHRNA5* on TC and LDL-cholesterol in never smokers, although this could have been attributable to type 1 error (Figure 6B, Table S12). There was very little evidence of paternal smoking effects on any outcome (Table S13). As with all stratified analyses, the potential for collider bias should be considered when interpreting these results.

**Figure 6.**
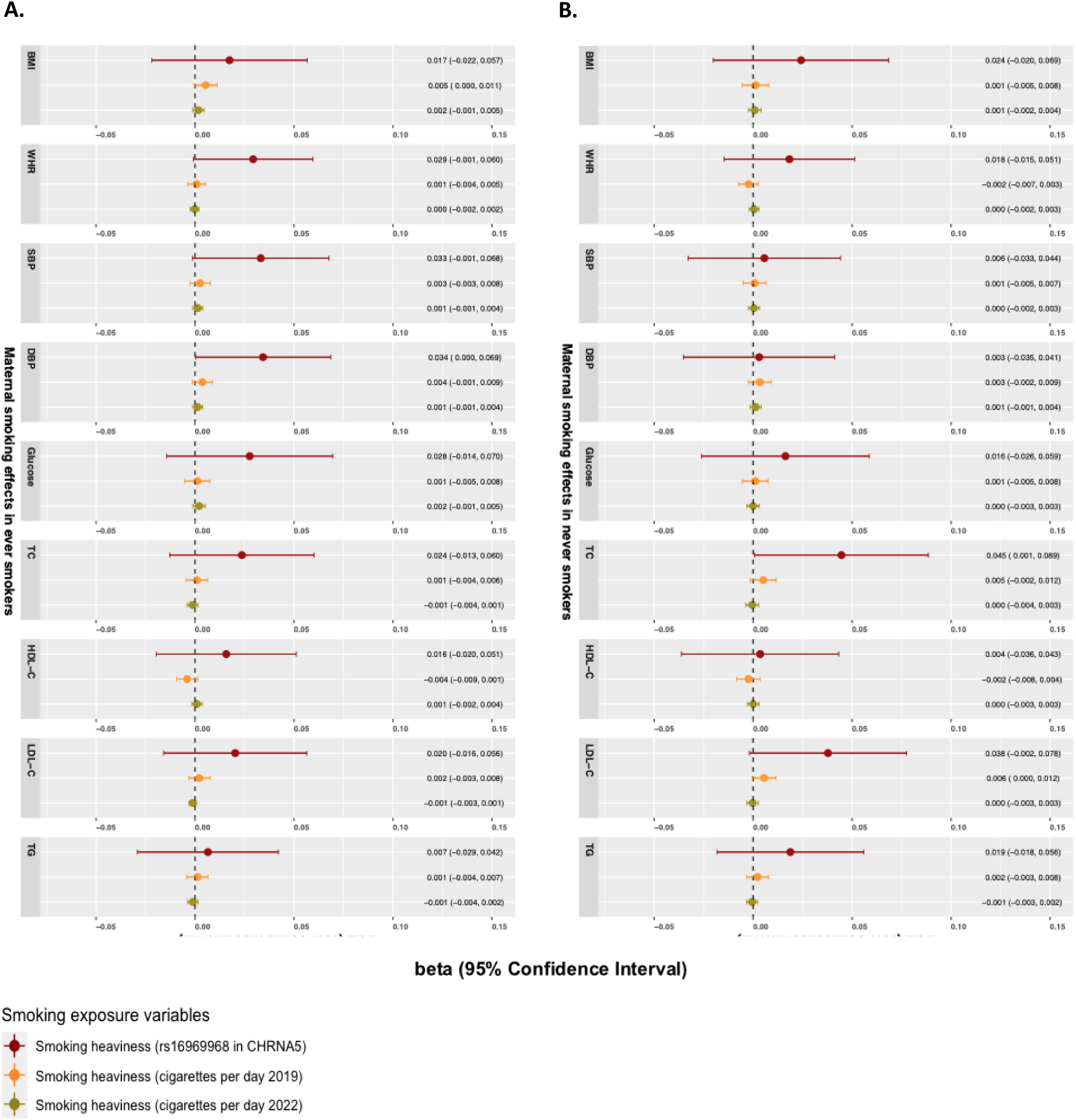
One-sample MR for maternal smoking heaviness onto 9 long term cardiometabolic outcomes in offspring in maternal ever and never smokers. A. Maternal smoking on offspring cardiometabolic risk factors in adulthood in maternal ever smoking group. B. Maternal smoking on offspring cardiometabolic risk factors in adulthood in maternal never smokers. Estimates represent the average change in offspring cardiometabolic outcomes per additional effect allele in the unweighted smoking GRS and per copy of the CHRNA5 risk allele, expressed in standard deviation (SD) units of each outcome. These estimates reflect associations between the parental genetic instruments and offspring outcomes, rather than causal effects per unit change in smoking heaviness.

## Discussion

Employing intergenerational MR analyses across three large cohorts, we found robust evidence supporting a causal effect of greater maternal smoking heaviness (but not initiation) on higher WHR in adult offspring. We also found suggestive evidence that greater maternal smoking heaviness causes greater offspring adult BMI and circulating CRP. We did not find robust evidence of maternal smoking effects on other cardiometabolic risk factors or of paternal smoking effects on any of the outcomes investigated. The MR models in this study cannot distinguish whether the estimated effects reflect maternal or paternal smoking before conception, during pregnancy, or after birth. They are therefore not specific to exposure in pregnancy, and we cannot determine from these results whether the observed effect of maternal smoking on adult offspring WHR is driven by pre-conception, intrauterine or postnatal mechanisms. A purely postnatal explanation appears less likely based on the weaker paternal effects observed, although this should be interpreted with caution, as unmeasured differences in parental caregiving, contact, or social expectations may contribute to stronger mother-offspring effects. Thus, our findings may reflect true effects that are similar across exposure to maternal smoking at different life stages, or alternatively we may have missed effects that are stronger or weaker at different critical or sensitive periods. If the assumptions underpinning our analyses hold, this study is consistent with previous non-genetic epidemiological findings, which identify an association between maternal smoking and higher central adiposity in offspring, that persists into adulthood (13, 14). Although there was some indication of an effect of maternal smoking on BMI and circulating CRP in the two-sample analyses, the BMI effect was weaker than that observed for WHR and did not replicate in the one-sample MR. CRP was not available for one-sample analysis. Overall, this pattern may suggest that any influence of maternal smoking heaviness relates specifically to the fat mass component of BMI.

Several potential mechanisms for the effect of maternal smoking on offspring adiposity have been proposed. In animal studies, maternal nicotine exposure has been associated with alterations in adipose tissue and glucose metabolism (82, 83). Although reduced fetal growth has been suggested as a DOHaD-related mechanism (84, 85), evidence from MR studies provides limited support for broad intrauterine programming effects (57), and such interpretations should therefore be made cautiously. Observational studies also show that associations between parental smoking and offspring overweight are often independent of fetal or postnatal growth and birth weight, which further limits support for a fetal growth–mediated explanation (12). Furthermore, sibling studies have reported that the association between parental smoking and offspring obesity/overweight in childhood and adulthood attenuates when assessed within siblings (20, 21). Such attenuation could indicate that familial confounding is an important driver of these associations, although the sibling estimates typically have wide confidence intervals which would also be consistent with meaningful causal effects. It is also difficult to directly compare effect sizes from our MR analyses with those from previous sibling studies, due to differences in the definition and measurement units of the smoking exposures and in estimands obtained across methods.

The limited evidence we observed for an effect of paternal smoking on WHR in adult offspring may be considered in the context of previous maternal and paternal comparison studies (22, 86). Such studies are often interpreted under the assumption that, if an intrauterine effect exists, the association of maternal smoking with offspring outcomes should be stronger than the equivalent paternal association (87). Earlier observational investigations and meta-analyses appeared consistent with this assumption, reporting more adverse maternal relative to paternal associations of parental smoking with childhood overweight and obesity (22, 86, 88). However, maternal-paternal comparisons are generally more informative for pregnancy outcomes, which occur during the exposure window itself, than for long-term offspring outcomes (23), where postnatal environmental and familial influences can accumulate and obscure intrauterine effects. Consistent with this, a more recent individual participant data meta-analysis including 229,158 families (24) reported maternal and paternal associations of comparable magnitude for childhood overweight, indicating that earlier maternal-paternal differences may have reflected residual confounding rather than true intrauterine effects. These limitations underscore the value of approaches such as MR in differentiating putative intrauterine causal effects from shared familial confounding when interrogating long-term offspring phenotypes.

Single-trait assortative mating, or nonrandom matching between spouses, occurs when partners are selected on the basis of a single phenotype (89). For instance, if individuals who smoke are more likely to select partners who also smoke, the resulting enrichment of smoking-concordant couples could bias associations in our study. In using a WLM to estimate parental genetic effects, we assume that the same phenotype was measured in the samples of mothers, fathers, and offspring, and that cross-locus genetic correlations due to assortative mating are absent. In addition, within a family context, it is difficult to fully unpick the effects of individual behaviour. Whilst it has been shown that the DNA methylation signature associated with passive smoke exposure is much less pronounced than that of own smoking, passive smoking or offspring’s own smoking behaviour may still have an impact on findings. Therefore, the effects that we found may encompass (i) maternal exposure to second-hand smoke from the partner during pregnancy, due to unmodelled assortative mating effects, (ii) postnatal exposure to second-hand smoke during the offspring’s childhood, and (iii) offspring smoking in adolescence and early adulthood.

There are several strengths of this study. For the main analysis, we took advantage of novel statistical methods (28, 60, 90, 91) recently developed to conduct intergenerational two-sample MR, by estimating the effects of parental genotype on offspring outcomes in adulthood, adjusting for the offspring’s own genotype and (when necessary) the other parent’s genotype. This approach removed the need to analyse a subset of individuals with complete maternal, paternal, and offspring genotype data available (as we have done for the one-sample MR analyses). In addition, we applied linear mixed model association methods to data from large population-based cohorts that comprised related individuals: HUNT, UKB and ALSPAC, increasing our available sample size considerably. We adjusted for the offspring’s and other parent’s genotype, which is crucial to preserve the key exclusion restriction assumption of MR (92). Importantly, we included birth weight as a positive control outcome, demonstrating that our method is able to detect the expected causal effect of maternal smoking on offspring birth weight. In particular, our MR results indicated an effect of both maternal and paternal smoking initiation on offspring birth weight. However, the presence of a paternal effect raises the possibility that the smoking initiation instrument may be picking up effects of other exposures besides smoking, via horizontal pleiotropy. We were also able to stratify MR analyses by parental smoking status; if parental smoking heaviness exhibits a putative causal effect in parental never smokers, this indicates our method is biased. We found some evidence that heavier maternal smoking causes lower offspring birth weight in the unstratified sample, but these effects were stronger in ever smokers.

An important limitation of this study is that despite relatively large sample sizes, low power was an issue, particularly within some of the stratified sensitivity analyses. These analyses should therefore be interpreted with appropriate caution. Moreover, smoking initiation is an example of a complex behavioural exposure where pleiotropy may be operating (93, 94). Smoking initiation has been shown to associate with risk-taking behaviours, personality traits, and externalising disorders (70, 71). To strengthen this study, we explored several indicators for smoking and conducted multiple sensitivity analyses. The relationship between smoking and body composition is particularly complex (68, 69). Suggestions of a common biological basis for nicotine addiction and obesity have been proposed, with higher adiposity influencing smoking behaviour (69, 95). This link identified between obesity and smoking behaviour may have implications for weight control and smoking prevention strategies (69). We identified several body composition measures which exhibit high genetic correlation with smoking indicators in publicly available GWAS data (52). Our MVMR analyses suggest that smoking initiation may be an unreliable exposure for MR analyses. In addition, a potential source of bias could arise from gene-sex interactions in the two-sample WLM analyses, if residual sex differences in SNP-outcome effects remain even after adjusting for offspring sex. This consideration is particularly relevant for WHR, which has well-established sex-specific genetic architecture (96), and in theory this could contribute to the stronger maternal MR estimate observed for this trait. However, since we observed similar maternal effects on BMI and CRP, which are less sexually dimorphic genetically, this is unlikely to be a major source of bias for our MR analyses. While all our outcome GWAS adjusted for offspring sex, they did not explicitly model gene-sex interactions, so some residual bias cannot be ruled out. Despite this, previously findings suggest that maternal and paternal BMI and glucose traits both exhibit null MR effects on offspring cardiometabolic risk factors in this data set (41, 42). Such bias is therefore unlikely to fully explain these patterns of associations. Finally, our study included only participants of White European ancestry from high income countries, therefore our results may not be generalisable to other populations.

In conclusion, our study provides evidence that heavier maternal smoking causes greater WHR in adult offspring, with more tentative indications of effects on offspring BMI and circulating CRP. We did not find robust evidence for maternal smoking effects on other cardiometabolic risk factors, or for effects of paternal smoking on any of the cardiometabolic factors examined. Triangulation with complementary study designs will be important for strengthening causal inference, and our findings, which are compatible with causal effects of maternal smoking on central adiposity in adult offspring, reinforce the public-health rationale for promoting smoking cessation prior to pregnancy.

## Acknowledgements

The Trøndelag Health Study (HUNT) is a collaboration between HUNT Research Centre (Faculty of Medicine and Health Sciences, Norwegian University of Science and Technology NTNU), Trøndelag County Council, Central Norway Regional Health Authority, and the Norwegian Institute of Public Health. In addition, we are extremely grateful to all the families who took part in the ALSPAC study, the midwives for their help in recruiting them, and the whole ALSPAC team, which includes interviewers, computer and laboratory technicians, clerical workers, research scientists, volunteers, managers, receptionists, and nurses. We would like to thank the UK Biobank study and all participants who contributed to it, as well as the authors of the GWAS who made their summary statistics available for the benefit of this work.

## Data and code sharing

Data access for the Avon Longitudinal Study of Parents and Children (ALSPAC) operates via a managed open access system. Approved proposals are reviewed by the ALSPAC Executive Committee. Full details are provided in the ALSPAC Data Management Plan (www.bristol.ac.uk/alspac/researchers/data-access/documents/alspac-data-management-plan.pdf). To request access to data from the Trøndelag Health Study (HUNT), researchers affiliated with Norwegian research institutes can apply for the use of HUNT data and biological samples, subject to approval by the Regional Committee for Medical and Health Research Ethics. Researchers from other countries may also apply if collaborating with a Norwegian Principal Investigator. All applications are reviewed by the HUNT Data Access Committee, and successful applicants are required to enter into data access and/or material transfer agreements. Detailed information on the application process, ethical requirements, and available datasets can be found on the HUNT website (www.ntnu.edu/hunt/data). Access to UK Biobank (UKB) data is obtained through a standard application process. Researchers register with UKB, submit a study proposal, and obtain approval from the UKB Access Management Team. Approved applicants receive access to de-identified data under UKB’s terms of use. Further information is available on the UK Biobank website (www.ukbiobank.ac.uk/enable-your-research/apply-for-access).

Upon publication, analysis code used in this study will be made publicly available via GitHub repository.

None of the material has been published or is under consideration for publication elsewhere.

