## Supplementary Materials - Text and Figures for "Effects of maternal and paternal smoking on offspring cardiometabolic risk factors in adulthood: a multi-method intergenerational Mendelian randomization study"

*Supplementary Material 1. STROBE-MR checklist of recommended items to address in reports of Mendelian randomization studies (1, 2).*

| Item No. | Section | | Checklist item | Relevant text from manuscript |
|---|---|---|---|---|
| 1 | **TITLE and ABSTRACT** | | Indicate Mendelian randomization (MR) as the study's design in the title and/or the abstract if that is a main purpose of the study | Title: Effects of maternal and paternal smoking on offspring cardiometabolic risk factors in adulthood: a multi-method intergenerational Mendelian randomisation study. |
| | **INTRODUCTION** | | | |
| 2 | **Background** | | Explain the scientific background and rationale for the reported study. What is the exposure? Is a potential causal relationship between exposure and outcome plausible? Justify why MR is a helpful method to address the study question | See entire Introduction and specifically:<br>We aimed to use intergenerational MR to test the hypothesis that maternal and paternal smoking – before, during, or after pregnancy – cause a more adverse offspring cardiometabolic risk factor profile in adulthood. |
| 3 | **Objectives** | | State specific objectives clearly, including pre-specified causal hypotheses (if any). State that MR is a method that, under specific assumptions, intends to estimate causal effects | See above and the following:<br>Mendelian randomization (MR) is a technique that, provided specific assumptions are met, exploits the random distribution of genetic variants from parents to offspring, independent of the influence of other traits. This reduces susceptibility to confounding factors, including confounding by undiagnosed existing disease (reverse causation) (24, 25). Recent developments in intergenerational MR methodology (26) enable indirect estimation of the effects of parental exposures on offspring adulthood outcomes by accounting for the direct fetal genotype contribution (6). |
| | **METHODS** | | | |
| 4 | **Study design and data sources** | | Present key elements of the study design early in the article. Consider including a table listing sources of data for all phases of the study. For each data source contributing to the analysis, describe the following: | See entire 'Data sources and instruments' section in 'Materials and methods', specifically:<br>Inclusion criteria and participating cohorts<br>We analysed three large population-based prospective cohorts that had genome-wide SNP data available in mothers, fathers and offspring and cardiometabolic risk factors measured in adult offspring: the Trøndelag Health Study (HUNT), the UK Biobank (UKB), and the Avon Longitudinal Study of Parents and Children (ALSPAC). The selection of study participants for our main two-sample MR analysis is presented in Figure 1. Study participant selection for one-sample MR is presented in Figure S1. |
| | | a) | Setting: Describe the study design and the underlying population, if possible. Describe the setting, locations, and relevant dates, including periods of recruitment, exposure, follow-up, and data collection, when available. | See entire 'Data sources and instruments' section and Figure 1, Supplementary Material 1, and Figure S1. |

| | | | | |
|---|---|---|---|---|
| | | b) | Participants: Give the eligibility criteria, and the sources and methods of selection of participants. Report the sample size, and whether any power or sample size calculations were carried out prior to the main analysis | See entire 'Data sources and instruments' section and Figure 1, Table 1, Supplementary Material, Figure S1. |
| | | c) | Describe measurement, quality control and selection of genetic variants | See entire 'Materials and methods' section, specifically the 'Genotyping, quality control and imputation in each cohort' section and Supplementary Material 1 |
| | | d) | For each exposure, outcome, and other relevant variables, describe methods of assessment and diagnostic criteria for diseases | See 'Offspring cardiometabolic risk factors and birthweight' and 'Parental smoking exposures' sections. |
| | | e) | Provide details of ethics committee approval and participant informed consent, if relevant | Informed consent was obtained from all participants and ethical approval was obtained from the Regional Committee for Medical and Health Research Ethics, Central Norway (REK Central application number 2018/2488) (HUNT), the North West Multi-centre Research Ethics Committee (MREC) (ref 11/NW/0382) (UKB), and the ALSPAC Ethics and Law Committee and the Local Research Ethics Committees (ALSPAC). |
| 5 | **Assumptions** | | Explicitly state the three core IV assumptions for the main analysis (relevance, independence and exclusion restriction) as well assumptions for any additional or sensitivity analysis | When genetic variants are used as instrumental variables in MR, the assumptions of instrumental variables must be met, i.e., the genetic variants used must (i) be strongly associated with the exposure of interest and relevant to the population studied, here pregnant women ("relevance"), (ii) not share common causes with the outcome ("independence"), and (iii) not affect the outcome other than through the exposure ("exclusion-restriction") (3).<br><br>To block the path from parental genotype through offspring genotype to the offspring outcome in an intergenerational context (which would be a violation of the "exclusion restriction" assumption), MR is robust if (i) summary outcome associations are extracted from adjusted GWAS (parental genotype adjusted for offspring genotype and if relevant, the other parent's genotype) in a two-sample setting, or (ii) the association between maternal or paternal unweighted GRS and offspring outcomes is adjusted for offspring genotype (and if relevant, other parent GRS), in a one-sample setting (4-7). There are other valid ways to conduct intergenerational one-sample MR (e.g. maternal non-transmitted alleles) (8), which will not be used in this current study. |
| 6 | **Statistical methods: main analysis** | | Describe statistical methods and statistics used | See entire 'Statistical analysis' section |
| | | a) | Describe how quantitative variables were handled in the analyses (i.e., scale, units, model) | See entire 'Materials and methods', specifically the 'Offspring cardiometabolic risk factors and birthweight' and 'Parental smoking exposures' sections, and 'Statistical analysis' section |
| | | b) | Describe how genetic variants were handled in the analyses and, if applicable, how their weights were selected | See 'Statistical analysis' and 'Parental smoking exposures' sections |

c) Describe the MR estimator (e.g. two-stage least squares, Wald ratio) and related statistics. Detail the included covariates and, in case of two-sample MR, whether the same covariate set was used for adjustment in the two samples

| | | | |
|---|---|---|---|
| | | | mean of the instrumental variable estimates (11). We checked for evidence of horizontal pleiotropy via Cochran's Q test for heterogeneity, and by testing whether the MR Egger intercept differed from zero. There is evidence of genetic variants associated with smoking also influencing body composition and possible bidirectional effects between smoking and body composition (13, 14). We therefore conducted the MR Steiger directionality test (15) in our two-sample analyses, to investigate the relationship of maternal and paternal smoking heaviness with WHR and BMI. This ensured the instruments used to estimate smoking influenced the smoking exposures first, rather than influencing smoking via BMI (reverse causality). We then applied multivariable MR (MVMR) to estimate the effect of smoking initiation and intensity on WHR, conditional on educational attainment and BMI. Educational attainment was included to capture socioeconomic correlations with smoking-associated variants, and BMI to account for shared genetic architecture with smoking (16-18). By modelling these exposures jointly, MVMR provides smoking-WHR estimates that account for these shared genetic pathways, under the multivariable IV assumptions (19, 20). Instrument strength in the multivariable MR MVMR models was evaluated using conditional F-statistics, which account for the inclusion of multiple, potentially correlated exposures. These analyses were performed using the TwoSampleMR R package (21). |
| 8 | **Sensitivity analyses and additional analyses** | Describe any sensitivity analyses or additional analyses performed (e.g. comparison of effect estimates from different approaches, independent replication, bias analytic techniques, validation of instruments, simulations) | See above as well as the 'Individual-level one-sample MR using GRS analysis' section. Also:<br><br>Stratified analysis<br><br>If our genetic instruments for smoking heaviness (including rs16969968 in the CHRNA5 gene) are valid, we would expect these variants to only indicate a causal effect when the mother/father is a smoker. If a causal effect is detected in parental never-smokers, this implies that the CHRNA5 gene is influencing the offspring outcome via mechanisms other than smoking heaviness (i.e. horizontal pleiotropy). We therefore repeated the CHRNA5 gene MR analyses, stratifying the sample to parental ever-smokers and never-smokers (47). For two sample analyses we only had a sufficiently large sample to run stratified analyses for birthweight (because the WLM relies on LD score regression (78), which has stringent sample size requirements), whereas we ran stratified one-sample analysis for adult outcomes too. |
| 9 | **Software and pre-registration** | | |
| | a) | Name statistical software and package(s), including version and settings used | These were generated using a weighted linear modelling (WLM) approach, implemented in the DONUTS R package, accounting for the correlation between the parental genotypes (21).<br><br>A linear mixed model using the fastGWA method (22) was used to account for cryptic relatedness. Parent-offspring pairs of European ancestry, for whom parental genotype and offspring phenotype data were available, were used. |

| | | | | |
|---|---|---|---|---|
| | | | | These analyses were conducted with HUNT, UKB and ALSPAC cohorts independently and then meta-analysed using a fixed effects model implemented in METAL version 2011-03-25 (23). Standard quality control (QC) procedures were carried out including calculating the Linkage Disequilibrium Score Regression (LDSC) intercept and attenuation ratio.<br><br>These analyses were performed using the TwoSampleMR R package (21).<br><br>A fixed-effects meta-analysis of regression coefficients from the three cohorts was conducted using Stata version 16 (24). |
| | | b) | State whether the study protocol and details were pre-registered (as well as when and where) | NA |
| | **RESULTS** | | | |
| 10 | **Descriptive data** | | | |
| | | a) | Report the numbers of individuals at each stage of included studies and reasons for exclusion. Consider use of a flow diagram | See Table 1, Figure 1, Figure S1, Table S4, Table S11, and Table S17, and:<br><br>Summary level two-sample Mendelian randomisation<br><br>The GWAS meta-analyses used for the SNP-outcome association in the two-sample MR included between 14,481 (for HbA1c with paternal genotype) and 511,253 (for DBP with offspring genotype) participants (Table S4) (25, 26).<br><br>Individual-level one-sample MR using GRS analysis<br><br>The individual-level one-sample GRS analysis comprised between 16,942 and 17,484 mother-father-offspring trios (Table 1 and Table S11). These one-sample analyses represent reduced form associations between parental genetic scores and offspring outcomes.<br><br>Stratified analysis<br><br>For GWAS of birthweight stratified by parental smoking status, analysis comprised between 10,719 and 136,933 participants in maternal ever smokers, 8,094 and 104,002 participants in maternal never smokers, 9,425 and 27,726 participants in paternal ever smokers and 5,367 and 18,718 participants in paternal never smokers (Table S17).<br><br>And:<br><br>The one-sample MR stratified smoking samples comprised between 8,814 and 9,065 participants in maternal ever smokers, 7,357 and 7,624 in maternal never smokers, 12,004 and 12,339 in paternal ever smokers and 4,445 and 4,623 in paternal never smokers, with numbers differing across outcomes (Table S11). |
| | | b) | Report summary statistics for phenotypic exposure(s), outcome(s), and other relevant variables (e.g. means, SDs, proportions) | See 'Summary level two-sample Mendelian randomisation' section and Supplementary |

| | | | | |
|---|---|---|---|---|
| | | c) | If the data sources include meta-analyses of previous studies, provide the assessments of heterogeneity across these studies | In addition, Cochran's Q test provided little evidence for between-SNP heterogeneity, and there was not strong evidence that the MR Egger intercept differed from zero (Table S7). |
| | | d) | For two-sample MR:<br>i. Provide justification of the similarity of the genetic variant-exposure associations between the exposure and outcome samples<br>ii. Provide information on the number of individuals who overlap between the exposure and outcome studies | The GWAS selected were conducted on participants of European ancestry to mitigate bias by population stratification.<br>Parent-offspring pairs of European ancestry, for whom parental genotype and offspring phenotype data were available, were used. These analyses were conducted with HUNT, UKB and ALSPAC cohorts independently and then meta-analysed using a fixed effects model implemented in METAL version 2011-03-25 (23). |
| 11 | **Main results** | | | |
| | | a) | Report the associations between genetic variant and exposure, and between genetic variant and outcome, preferably on an interpretable scale | See 'Parental smoking exposures' section. |
| | | b) | Report MR estimates of the relationship between exposure and outcome, and the measures of uncertainty from the MR analysis, on an interpretable scale, such as odds ratio or relative risk per SD difference | See Results section, specifically:<br><br>We found strong evidence of an average change in adult offspring WHR per one standard deviation (SD) increase in maternal cigarettes smoked per day of 0.25 SD (95% CI: 0.13, 0.38) using GWAS carried out in 2019, 0.19 SD (95% CI: 0.08, 0.31) using the GWAS carried out in 2022, and 0.27 SD (95% CI: 0.07, 0.47) using CHRNA5 rs16969968 (Figure 3A, Table S5). There was weaker evidence that increased maternal smoking heaviness had a causal effect on higher BMI (average change in SD of BMI in adult offspring per maternal cigarettes smoked per day of 0.23 SD (95% CI: 0.07, 0.39) using GWAS carried out in 2019, 0.14 SD (95% CI: 0.00, 0.29) using GWAS carried out in 2022 and 0.11 SD (95% CI: -0.13, 0.35) using CHRNA5 rs16969968) and CRP (average change in SD of CRP in adult offspring 1SD greater maternal cigarettes smoked per day of 0.24 SD (95% CI: 0.08, 0.40) using GWAS carried out in 2019, 0.20 SD (95% CI: 0.04, 0.36) using GWAS carried out in 2022, and 0.25 SD (95% CI: 0.01, 0.49) using CHRNA5 rs16969968). Corresponding paternal effect estimates are presented in Table S6. In the analyses using the trios WLM for the outcome GWAS (in which the parental genetic effects were adjusted for the other parent's genotype as well as the offspring's genotype), results were similar, although the confidence intervals were wider due to the lower statistical power of the trios WLM versus the duos WLM (Figure S2).<br><br>And:<br><br>The individual-level one-sample GRS analysis comprised between 16,942 and 17,484 mother-father-offspring trios (Table 1 and Table S11). These one-sample analyses represent reduced form associations between parental genetic scores and offspring outcomes. One-sample MR in the total population indicated that higher genetically predicted maternal smoking heaviness (using rs16969968 in the CHRNA5 SNP) had a positive effect on WHR, TC, and LDL-cholesterol in |

| | | | | |
|---|---|---|---|---|
| | | | | offspring in later life (Figure 4A, Table S12). There was no evidence of paternal smoking effects (Figure 4B, Table S13). Cohort-specific results are presented in Tables S14-S16. Effect estimates obtained using the unweighted smoking GRS were smaller than those from rs16969968 in CHRNA5, consistent with differences in instrument scale: a single CHRNA5 allele has a larger established effect on smoking intensity than a one-allele change in the unweighted multi-locus score. Consequently, the unweighted GRS yields more diluted reduced-form associations. |
| | | c) | If relevant, consider translating estimates of relative risk into absolute risk for a meaningful time period | NA |
| | | d) | Consider plots to visualize results (e.g. forest plot, scatterplot of associations between genetic variants and outcome versus between genetic variants and exposure) | See entire 'Results' section as well as Supplementary Figures. |
| 12 | **Assessment of assumptions** | | | |
| | | a) | Report the assessment of the validity of the assumptions | Point estimates from the two-sample weighted median, weighted mode and MR-Egger methods were similar to IVW estimates and evidence remained robust for the maternal effect for smoking heaviness on offspring WHR in later life (Table S5). In addition, Cochran's Q test provided little evidence for between-SNP heterogeneity, and there was not strong evidence that the MR Egger intercept differed from zero (Table S7). This suggests little evidence that horizontal pleiotropy is driving these results. After correcting for multiple testing using a Bonferroni test, evidence of an effect remained for maternal smoking heaviness on increased WHR and BMI only (using p=0.004). Findings for adult offspring outcomes using the unadjusted GWAS sensitivity analyses are presented in Table S8 for maternal exposures and Table S9 for paternal exposures. MVMR analyses indicated that there remained strong evidence for the effect of maternal smoking heaviness on WHR in offspring in later life, after accounting for BMI as well as educational attainment (Table S10). There was no evidence of an effect of either educational attainment or BMI on WHR in offspring, after accounting for any maternal smoking indicator. These results should be interpreted with caution, however, since conditional F-statistic was <10 for each of the smoking exposures, indicating potential for weak instrument bias. |
| | | b) | Report any additional statistics (e.g., assessments of heterogeneity across genetic variants, such as $I^2$, Q statistic or E-value) | In addition, Cochran's Q test provided little evidence for between-SNP heterogeneity, and there was not strong evidence that the MR Egger intercept differed from zero (Table S7). |
| 13 | **Sensitivity analyses and additional analyses** | | | |
| | | a) | Report any sensitivity analyses to assess the robustness of the main results to violations of the assumptions | Point estimates from the two-sample weighted median, weighted mode and MR-Egger methods were similar to IVW estimates and evidence remained robust for the maternal effect for smoking heaviness on offspring WHR in later |

life (Table S5). In addition, Cochran's Q test provided little evidence for between-SNP heterogeneity, and there was not strong evidence that the MR Egger intercept differed from zero (Table S7). This suggests little evidence that horizontal pleiotropy is driving these results. After correcting for multiple testing using a Bonferroni test, evidence of an effect remained for maternal smoking heaviness on increased WHR and BMI only (using p=0.004). Findings for adult offspring outcomes using the unadjusted GWAS sensitivity analyses are presented in Table S8 for maternal exposures and Table S9 for paternal exposures. MVMR analyses indicated that there remained strong evidence for the effect of maternal smoking heaviness on WHR in offspring in later life, after accounting for BMI as well as educational attainment (Table S10). There was no evidence of an effect of either educational attainment or BMI on WHR in offspring, after accounting for any maternal smoking indicator. These results should be interpreted with caution, however, since conditional F-statistic was <10 for each of the smoking exposures, indicating potential for weak instrument bias.

| | | | |
|---|---|---|---|
| | | | group the effects of maternal smoking heaviness flipped sign and became weakly positive. Taken together, these observations support our use of birth weight as a positive control outcome. In the analyses using the adjusted GWAS, the direction of effect was often consistent with analyses using the unadjusted GWAS, but the adjusted GWAS had lower statistical power, resulting in weaker evidence (Figure S3, Figure S4). There was little evidence that paternal smoking heaviness caused offspring birth weight (Figure 5, Figure S3; Table S18).<br>The one-sample MR stratified smoking samples comprised between 8,814 and 9,065 participants in maternal ever smokers, 7,357 and 7,624 in maternal never smokers, 12,004 and 12,339 in paternal ever smokers and 4,445 and 4,623 in paternal never smokers, with numbers differing across outcomes (Table S11). Whilst the sample size was smaller and thus power was low, the magnitude of effect became slightly stronger in ever smokers for the effects of maternal smoking heaviness instrumented by rs16969968 in CHRNA5 on WHR and maternal smoking heaviness instrumented by cigarettes per day on BMI (Figure 6A, Table S12). There was additionally some evidence of an effect of maternal smoking heaviness instrumented by rs16969968 in CHRNA5 on DBP and SBP in ever smokers. There was weak evidence of an effect of maternal smoking heaviness instrumented by rs16969968 in CHRNA5 on TC and LDL-cholesterol in never smokers, although this could have been attributable to type 1 error (Figure 6B, Table S12). There was very little evidence of paternal smoking effects on any outcome (Table S13). |
| | | c) Report any assessment of direction of causal relationship (e.g., bidirectional MR) | NA |
| | | d) When relevant, report and compare with estimates from non-MR analyses | See Discussion section. |
| | | e) Consider additional plots to visualize results (e.g., leave-one-out analyses) | See entire ‘Results’ section. |
| | **DISCUSSION** | | |
| 14 | **Key results** | Summarize key results with reference to study objectives | Employing intergenerational MR analyses across three large cohorts, we found robust evidence supporting a causal effect of greater maternal smoking heaviness (but not initiation) on higher WHR in adult offspring. We also found suggestive evidence that greater maternal smoking heaviness causes greater offspring adult BMI and circulating CRP. We did not find robust evidence of maternal smoking effects on other cardiometabolic risk factors or of paternal smoking effects on any of the outcomes investigated. The MR models presented in this study test an omnibus exposure that may capture the potential effects of maternal and paternal smoking before, during, and after pregnancy. They are therefore not specific to exposure in pregnancy, and we cannot determine from these results whether the observed effect of maternal smoking on adult offspring WHR is driven by pre-conception, intrauterine or postnatal mechanisms. Despite this, a purely postnatal explanation may be less likely given the weaker paternal associations that we observed. Thus, our findings may reflect true effects that are similar across exposure to maternal smoking at different life stages, or alternatively we may have missed effects that are stronger or weaker at different critical or sensitive periods. If the assumptions |

| | | | |
|---|---|---|---|
| | | | underpinning our analyses hold, this study is consistent with previous non-genetic epidemiological findings, which identify an association between maternal smoking and higher central adiposity in offspring, that persists into adulthood (14, 15). Although there was some indication of an effect of maternal smoking on BMI and circulating CRP in the two-sample analyses, the BMI effect was weaker than that observed for WHR and did not replicate in the one-sample MR. CRP was not available for one-sample analysis. Overall, this pattern may suggest that any influence of maternal smoking heaviness relates specifically to the fat mass component of BMI. |
| 15 | **Limitations** | Discuss limitations of the study, taking into account the validity of the IV assumptions, other sources of potential bias, and imprecision. Discuss both direction and magnitude of any potential bias and any efforts to address them | An important limitation of this study is that despite relatively large sample sizes, low power was an issue, particularly within some of the stratified sensitivity analyses. These analyses should therefore be interpreted with appropriate caution. Moreover, smoking initiation is an example of a complex behavioural exposure where pleiotropy may be operating (92, 93). Smoking initiation has been shown to associate with risk-taking behaviours, personality traits, and externalising disorders (69, 70). To strengthen this study, we explored several indicators for smoking and conducted multiple sensitivity analyses. The relationship between smoking and body composition is particularly complex (66, 67). Suggestions of a common biological basis for nicotine addiction and obesity have been proposed, with higher adiposity influencing smoking behaviour (67, 94). This link identified between obesity and smoking behaviour may have implications for weight control and smoking prevention strategies (67). We identified several body composition measures which exhibit high genetic correlation with smoking indicators in publicly available GWAS data (50). Our MVMR analyses suggest that smoking initiation may be an unreliable exposure for MR analyses. In addition, a potential source of bias could arise from gene-sex interactions in the two-sample WLM analyses, if residual sex differences in SNP-outcome effects remain even after adjusting for offspring sex. This consideration is particularly relevant for WHR, which has well-established sex-specific genetic architecture (95), and in theory this could contribute to the stronger maternal MR estimate observed for this trait. However, since we observed similar maternal effects on BMI and CRP, which are less sexually dimorphic genetically, this is unlikely to be a major source of bias for our MR analyses. While all our outcome GWAS adjusted for offspring sex, they did not explicitly model gene-sex interactions, so some residual bias cannot be ruled out. Despite this, previously findings suggest that maternal and paternal BMI and glucose traits both exhibit null MR effects on offspring cardiometabolic risk factors in this data set (39, 40). Such bias is therefore unlikely to fully explain these patterns of associations. Finally, our study included only participants of White European ancestry from high income countries, therefore our results may not be generalisable to other populations. |
| 16 | **Interpretation** | | |
| | a) | Meaning: Give a cautious overall interpretation of results in the context of their limitations and in comparison with other studies | See entire Discussion section. |

| | | |
|---|---|---|
| | b) Mechanism: Discuss underlying biological mechanisms that could drive a potential causal relationship between the investigated exposure and the outcome, and whether the gene-environment equivalence assumption is reasonable. Use causal language carefully, clarifying that IV estimates may provide causal effects only under certain assumptions | Several potential mechanisms for the causal effect of maternal smoking on offspring adiposity have been previously suggested. In animal studies, an association between maternal nicotine exposure and alterations in adipose tissue and glucose metabolism has been reported (81, 82). This lends biological support to the hypothesis that maternal smoking during pregnancy may cause increased offspring adiposity. Research in human populations has suggested that a reduction in fetal growth may 'programme' the offspring early in life to later develop obesity (83, 84), consistent with the DOHaD hypothesis. However, observational associations of parental smoking with child overweight are often independent of fetal and postnatal growth and birth weight, arguing against this interpretation (12). Furthermore, sibling studies have reported that the association between parental smoking and offspring obesity/overweight in childhood and adulthood attenuates when assessed within siblings (20, 21). Such attenuation could indicate that familial confounding is an important driver of these associations, although the sibling estimates typically have wide confidence intervals which would also be consistent with meaningful causal effects. It is also difficult to directly compare effect sizes from our MR analyses with those from previous sibling studies, due to differences in the definition and measurement units of the smoking exposures and in estimands obtained across methods.<br>The limited evidence we observed for an effect of paternal smoking on WHR in adult offspring are supported by maternal and paternal comparison studies (22, 85). The assumption in these study designs is that if an intrauterine effect of the maternal exposure on the offspring outcome is present, the association of the maternal exposure with the outcome is expected to be stronger than the association of the equivalent paternal exposure (86). This is shown in several investigations of parental smoking on offspring outcomes (22, 85-87). For example, a meta-analysis comprising 12 observational studies published between 2008 and 2013 (n=109,838) indicated larger effect estimates for the association of maternal smoking in pregnancy with childhood overweight and obesity compared with paternal smoking in mutually adjusted models (22).<br>Single-trait assortative mating, or nonrandom matching between spouses, occurs when partners are selected on the basis of a single phenotype (88). For example, the parents included in our study may have more similar smoking habits, which could induce bias. In using a WLM to estimate parental genetic effects, we assume that the same phenotype was measured in the samples of mothers, fathers, and offspring, and that cross-locus genetic correlations due to assortative mating are absent. In addition, within a family context, it is difficult to fully unpick the effects of individual behaviour. Whilst it has been shown that the DNA methylation signature associated with passive smoke exposure is much less pronounced than that of own smoking, passive smoking or offspring's own smoking behaviour may still have an impact on findings. Therefore, the effects that we found may encompass (i) maternal exposure to second-hand smoke from the partner during pregnancy, due to unmodelled assortative mating effects, (ii) postnatal exposure to second-hand smoke during the offspring's childhood, and (iii) offspring smoking in adolescence and early adulthood. |

| | | | | |
|---|---|---|---|---|
| | | c) | Clinical relevance: Discuss whether the results have clinical or public policy relevance, and to what extent they inform effect sizes of possible interventions | In conclusion, our study provides evidence that heavier maternal smoking causes greater WHR in adult offspring, with more tentative indications of effects on offspring BMI and circulating CRP. We did not find robust evidence for maternal smoking effects on other cardiometabolic risk factors, or for effects of paternal smoking on any of the cardiometabolic factors examined. If replicated, our evidence for a causal association between maternal smoking and increased central adiposity in adult offspring provides further motivation for promoting smoking cessation prior to pregnancy. |
| 17 | **Generalizability** | | Discuss the generalizability of the study results (a) to other populations, (b) across other exposure periods/timings, and (c) across other levels of exposure | Finally, our study included only participants of White European ancestry from high income countries, therefore our results may not be generalisable to other populations. |
| | **OTHER INFORMATION** | | | |
| 18 | **Funding** | | Describe sources of funding and the role of funders in the present study and, if applicable, sources of funding for the databases and original study or studies on which the present study is based | Sources of funding.<br><br>The genotyping in HUNT was financed by the National Institutes of Health (NIH); University of Michigan; The Research Council of Norway; The Liaison Committee for Education, Research and Innovation in Central Norway; and the Joint Research Committee between St. Olavs hospital and the Faculty of Medicine and Health Sciences, NTNU. This work was in part supported by the Integrative Epidemiology Unit which receives funding from the UK Medical Research Council and the University of Bristol (MC_UU_00032/1 and MC_UU_00032/5). The UK Medical Research Council and Wellcome (Grant ref: MR/Z505924/1) and the University of Bristol provide core support for ALSPAC. Genotyping of the ALSPAC maternal samples was funded by the Wellcome Trust (WT088806) and the offspring samples were genotyped by Sample Logistics and Genotyping Facilities at the Wellcome Trust Sanger Institute and LabCorp (Laboratory Corporation of America) using support from 23andMe. A comprehensive list of grants funding is available on the ALSPAC website (http://www.bristol.ac.uk/alspac/external/documents/grant-acknowledgements.pdf); this research was specifically funded by Wellcome Trust and MRC (076467/Z/05/Z, 102215/2/13/2) and British Heart Foundation (BHF) (CS/15/6/31468). This publication is the work of the authors and will serve as guarantors for the contents of this paper. GDS conducts research at the NIHR Biomedical Research Centre at the University Hospitals Bristol NHS Foundation Trust and the University of Bristol. The views expressed in this publication are those of the author(s) and not necessarily those of the NHS, the National Institute for Health Research or the Department of Health. GMP was supported by the GW4 Biomed Doctoral Training Programme, awarded to the Universities of Bath, Bristol, Cardiff and Exeter from the Medical Research Council (MRC)/UKRI (MR/N0137941/1) whilst completing this work. GW and NMW are supported by a National Health and Medical Research Council (NHMRC; Australia) Emerging Leadership Fellowship (APP2008723). DME is funded by an |

| | | | |
|---|---|---|---|
| | | | Australian National Health and Medical Research Council Investigator Grant (APP2017942). GHM is the recipient of an Australian Research Council Discovery Early Career Award (Project number: DE220101226) funded by the Australian Government and supported by the Research Council of Norway (Project grant: 325640 & Mobility grant: 287198). DAL's contribution to this work is supported by the European Research council (101021566) and BHF (CH/F/20/90003 and AA/18/1/34219), and TAB's work was supported by the BHF (AA/18/1/34219). LB, BOA, and BMB work in a research unit funded by the Liaison Committee for education, research and innovation in Central Norway and the Joint Research Committee between St. Olavs Hospital and the Faculty of Medicine and Health Sciences, NTNU. |
| 19 | **Data and data sharing** | Provide the data used to perform all analyses or report where and how the data can be accessed and reference these sources in the article. Provide the statistical code needed to reproduce the results in the article, or report whether the code is publicly accessible and if so, where | Data access for the Avon Longitudinal Study of Parents and Children (ALSPAC) operates via a managed open access system. Approved proposals are reviewed by the ALSPAC Executive Committee. Full details are provided in the ALSPAC Data Management Plan (www.bristol.ac.uk/alspac/researchers/data-access/documents/alspac-data-management-plan.pdf). To request access to data from the Trøndelag Health Study (HUNT), researchers affiliated with Norwegian research institutes can apply for the use of HUNT data and biological samples, subject to approval by the Regional Committee for Medical and Health Research Ethics. Researchers from other countries may also apply if collaborating with a Norwegian Principal Investigator. All applications are reviewed by the HUNT Data Access Committee, and successful applicants are required to enter into data access and/or material transfer agreements. Detailed information on the application process, ethical requirements, and available datasets can be found on the HUNT website (www.ntnu.edu/hunt/data). Access to UK Biobank (UKB) data is obtained through a standard application process. Researchers register with UKB, submit a study proposal, and obtain approval from the UKB Access Management Team. Approved applicants receive access to de-identified data under UKB's terms of use. Further information is available on the UK Biobank website (www.ukbiobank.ac.uk/enable-your-research/apply-for-access)<br><br>None of the material has been published or is under consideration for publication elsewhere. |
| 20 | **Conflicts of Interest** | All authors should declare all potential conflicts of interest | All authors declare no competing interests. |

This checklist is copyrighted by the Equator Network under the Creative Commons Attribution 3.0 Unported (CC BY 3.0) license.

Supplementary Material 1. Further information on genotyping, quality control and imputation in each cohort

HUNT samples were genotyped using one of three different Illumina HumanCoreExome arrays (HumanCoreExome12 v1.0, HumanCoreExome12 v1.1, and UM HUNT Biobank v1.0) (27). Genomic position, strand orientation, and the reference allele of genotyped variants were determined by aligning their probe sequences against the human genome (Genome Reference Consortium Human genome build 37 and revised Cambridge Reference Sequence of the human mitochondrial DNA; http://genome.ucsc.edu) using BLAT (28). Ancestry of all samples was inferred by projecting all genotyped samples into the space of the principal components of the Human Genome Diversity Project (HGDP) reference panel (938 unrelated individuals; downloaded from http://csg.sph.umich.edu/chaolong/LASER/) (29, 30) using PLINK v1.9039(31). The resulting genotype data were phased using Eagle2 v2.340(32). Imputation was performed on the samples of recent European ancestry using Minimac3 (v2.0.1, http://genome.sph.umich.edu/wiki/Minimac3) (33) with default settings (2.5 Mb reference based chunking with 500 kb windows) and a customized Haplotype Reference consortium release 1.1 (HRC v1.1) for autosomal variants and HRC v1.1 for chromosome X variants (34). This process was explained previously (4).

UKB participants were assayed using two similar genotyping arrays (35). A subset of 49,950 participants involved in the UK Biobank Lung Exome Variant Evaluation (UK BiLEVE) study were genotyped at 807,411 markers using the Applied Biosystems UK BiLEVE Axiom Array by Affymetrix (now part of Thermo Fisher Scientific), which is described elsewhere6. Following this, 438,427 participants were genotyped using the closely related Applied Biosystems UK Biobank Axiom Array (825,927 markers) that shares 95% of marker content with the UK BiLEVE Axiom Array. The marker content of the UK Biobank Axiom array was chosen to capture genome-wide genetic variation (single nucleotide polymorphism (SNPs) and short insertions and deletions (indels)). Many markers were included because of known associations with, or possible roles in, disease. The array also includes coding variants across a range of minor allele frequencies (MAFs), including rare markers (<1% MAF); and markers that provide good genome-wide coverage for imputation in European populations in the common (>5%) and low frequency (1–5%) MAF ranges.

ALSPAC mothers were genotyped using Illumina human660K quad single nucleotide polymorphism (SNP) chip, and ALSPAC children were genotyped using Illumina HumanHap550 quad genome-wide SNP genotyping platform (36). ALSPAC fathers were genotypes using the 1000 Genomes phase 1 panel (37). Genotype data were imputed against the Haplotype Reference Consortium v1.1 reference panel, after performing the QC procedure (minor allele frequency (MAF)≥1%, a call rate≥95%, in Hardy–Weinberg equilibrium (HWE), correct sex assignment, no evidence of cryptic relatedness, and of European descent).

**Figure S1.** Study design overview and flowchart for participants included in one-sample MR analysis.

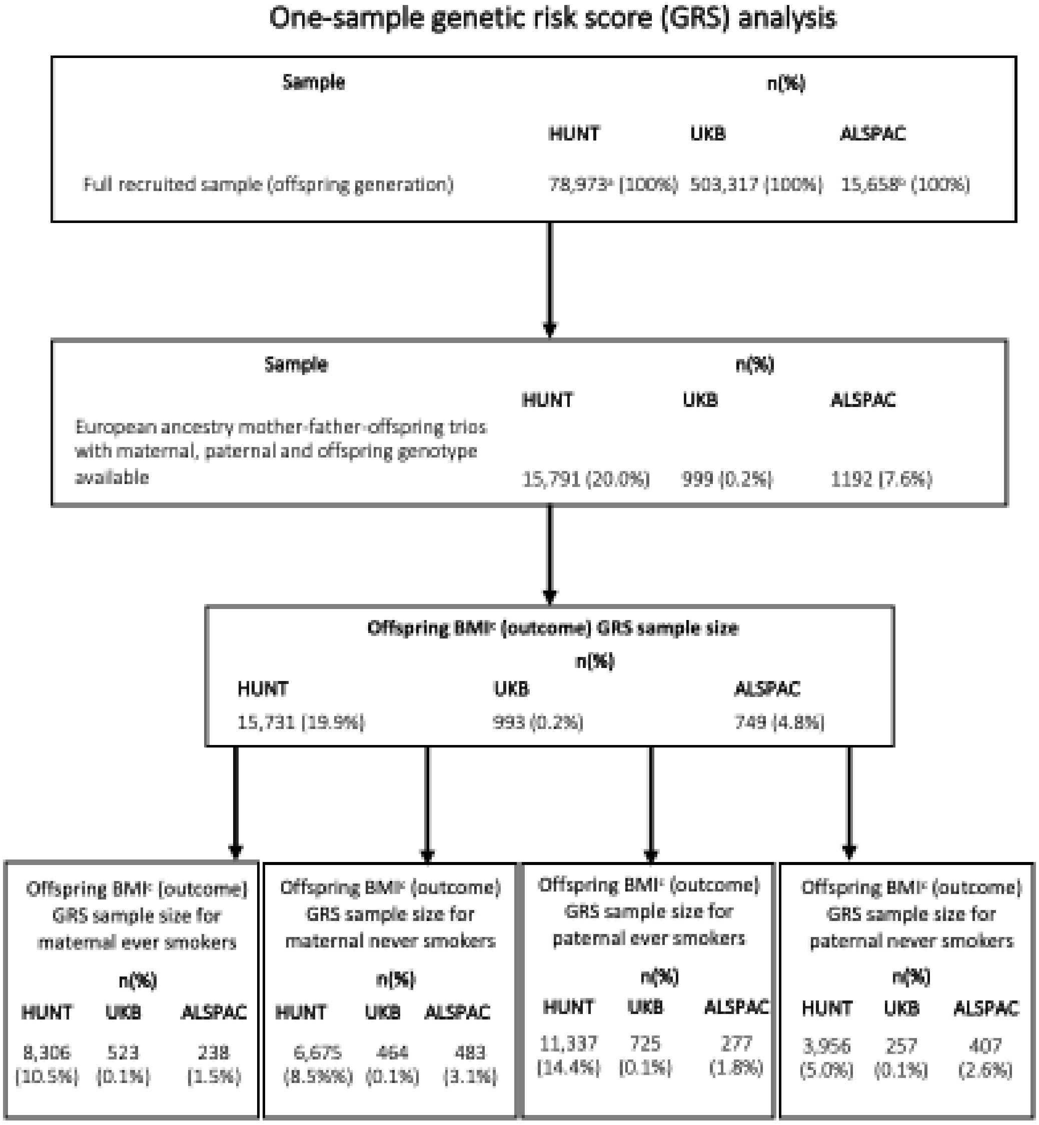


BMI: body mass index, HUNT: Trøndelag Health Study, UKB: UK Biobank, ALSPAC: Avon Longitudinal Study of Parents and Children, GRS: Genetic risk score. a: total number of participants in the HUNT2 and HUNT3 study waves, b: total number of fetuses included in ALSPAC (including participants recruited after 7 years of age), c: BMI was used as an example, samples were similar for other adult outcomes.

**Figure S2.** Two-sample MR for maternal and paternal smoking heaviness and initiation onto 11 long term cardiometabolic outcomes in offspring. Parental genotype was adjusted for offspring genotype and the other parent's genotype (trios WLM). A. Maternal smoking on offspring cardiometabolic risk factors in adulthood. B. Paternal smoking on offspring cardiometabolic risk factors in adulthood. MR estimates for the effect of smoking heaviness can be interpreted as the average change in the outcome, in standard deviation (SD) units, per one SD greater parental cigarettes smoked per day. MR estimates for the effect of smoking initiation can be interpreted as the average change in the outcome, in SD units, per doubling in the odds of smoking initiation.

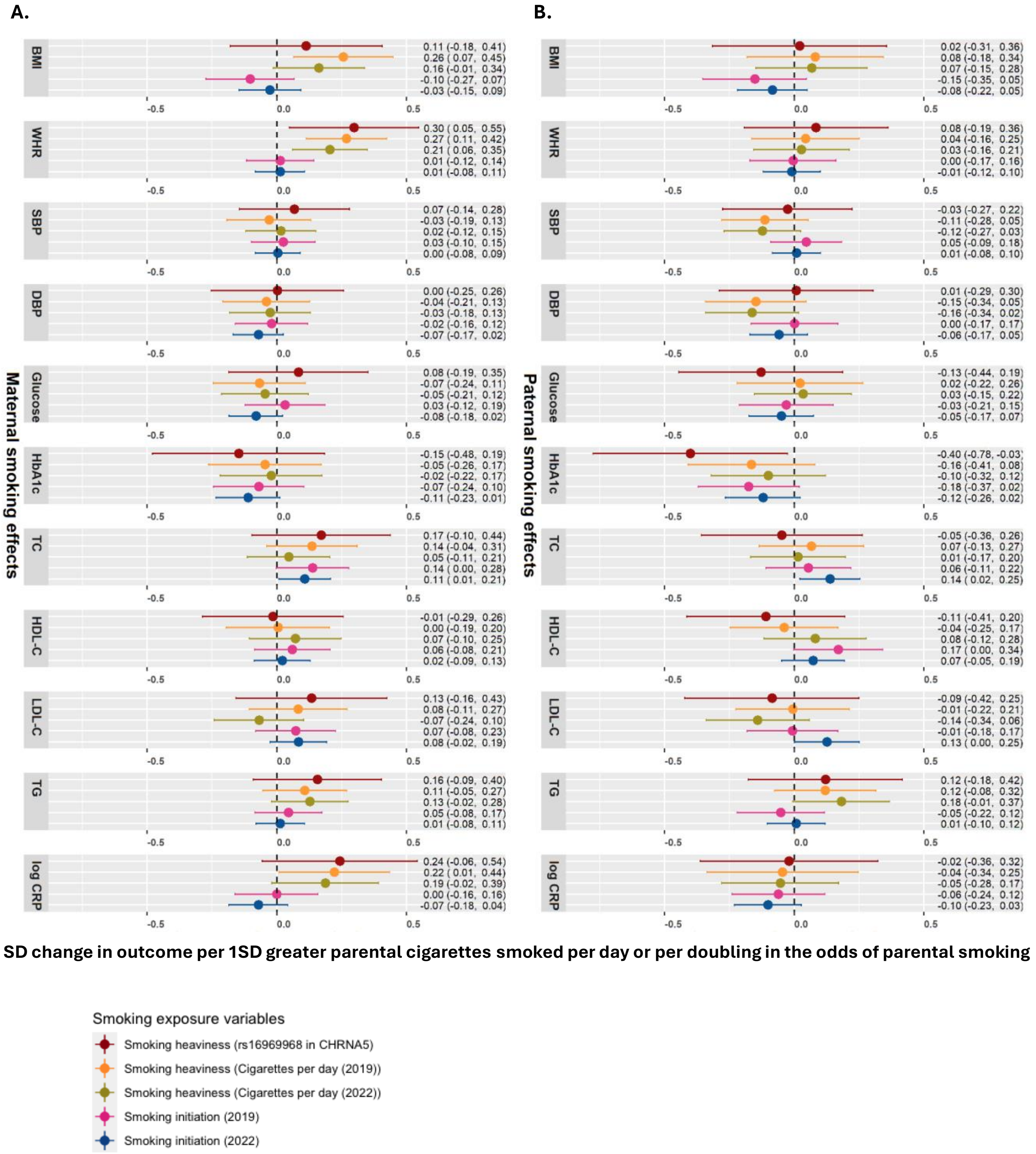

**Figure S3.** Two-sample MR for maternal and paternal smoking heaviness and initiation onto offspring birth weight, stratified by parental smoking status. Parental genotype was adjusted for offspring genotype and the other parent's genotype (trios WLM). A. Maternal smoking on offspring birth weight. B. Paternal smoking on offspring birth weight. MR estimates for the effect of smoking heaviness can be interpreted as the average change in the outcome, in standard deviation (SD) units, per one SD greater parental cigarettes smoked per day. MR estimates for the effect of smoking initiation can be interpreted as the average change in the outcome, in SD units, per doubling in the odds of smoking initiation.

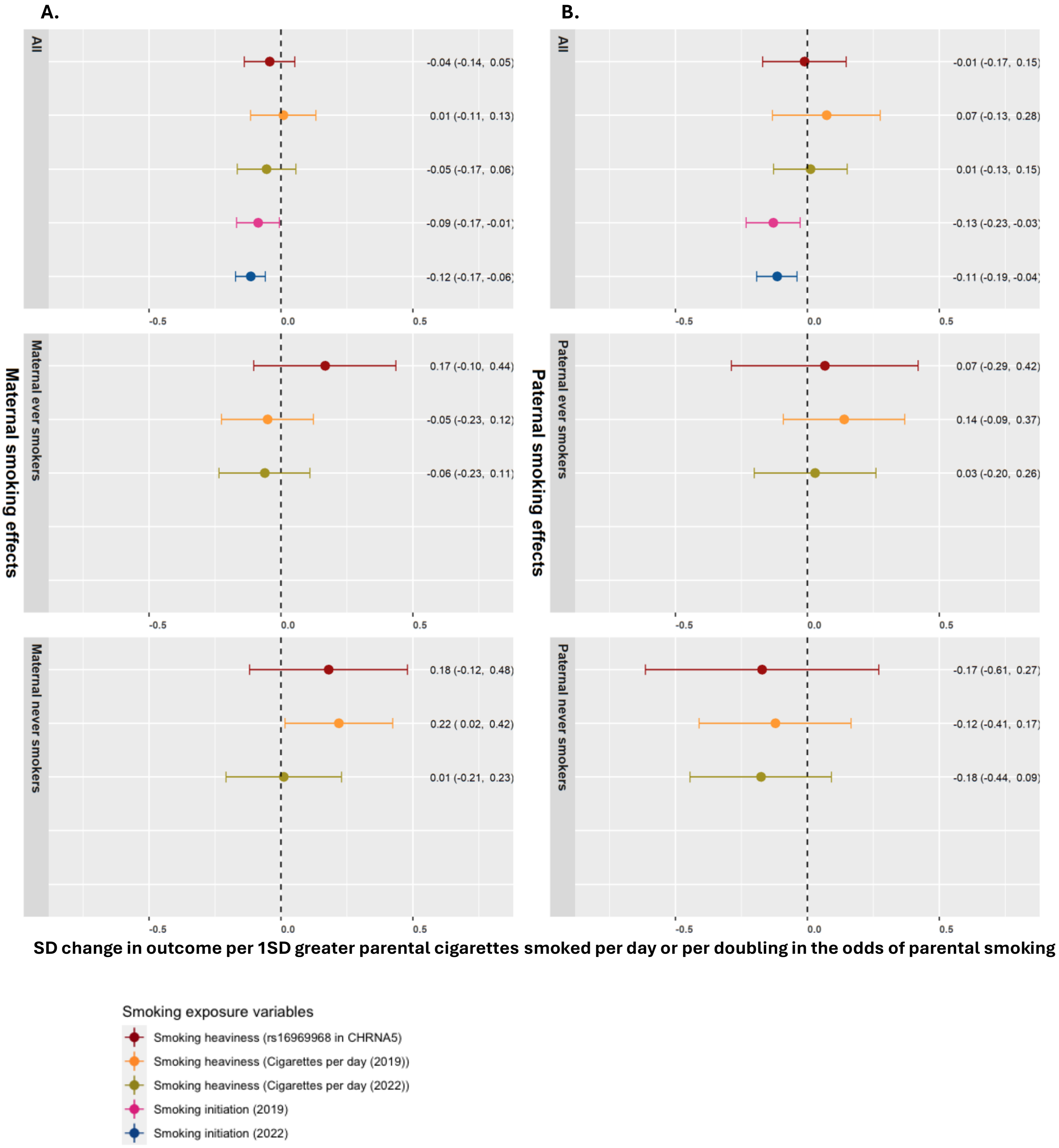

**Figure S4.** Two-sample MR for maternal smoking heaviness and initiation onto offspring birth weight, stratified by maternal smoking status. Maternal genotype was adjusted for only offspring genotype (duos WLM). MR estimates for the effect of smoking heaviness can be interpreted as the average change in the outcome, in standard deviation (SD) units, per one SD greater parental cigarettes smoked per day. MR estimates for the effect of smoking initiation can be interpreted as the average change in the outcome, in SD units, per doubling in the odds of smoking initiation. We did not conduct MR analyses using the duos WLM for paternal smoking exposures, since paternal duos WLM results would be affected by collider bias if there are maternal genetic effects (due to conditioning on offspring genotype)

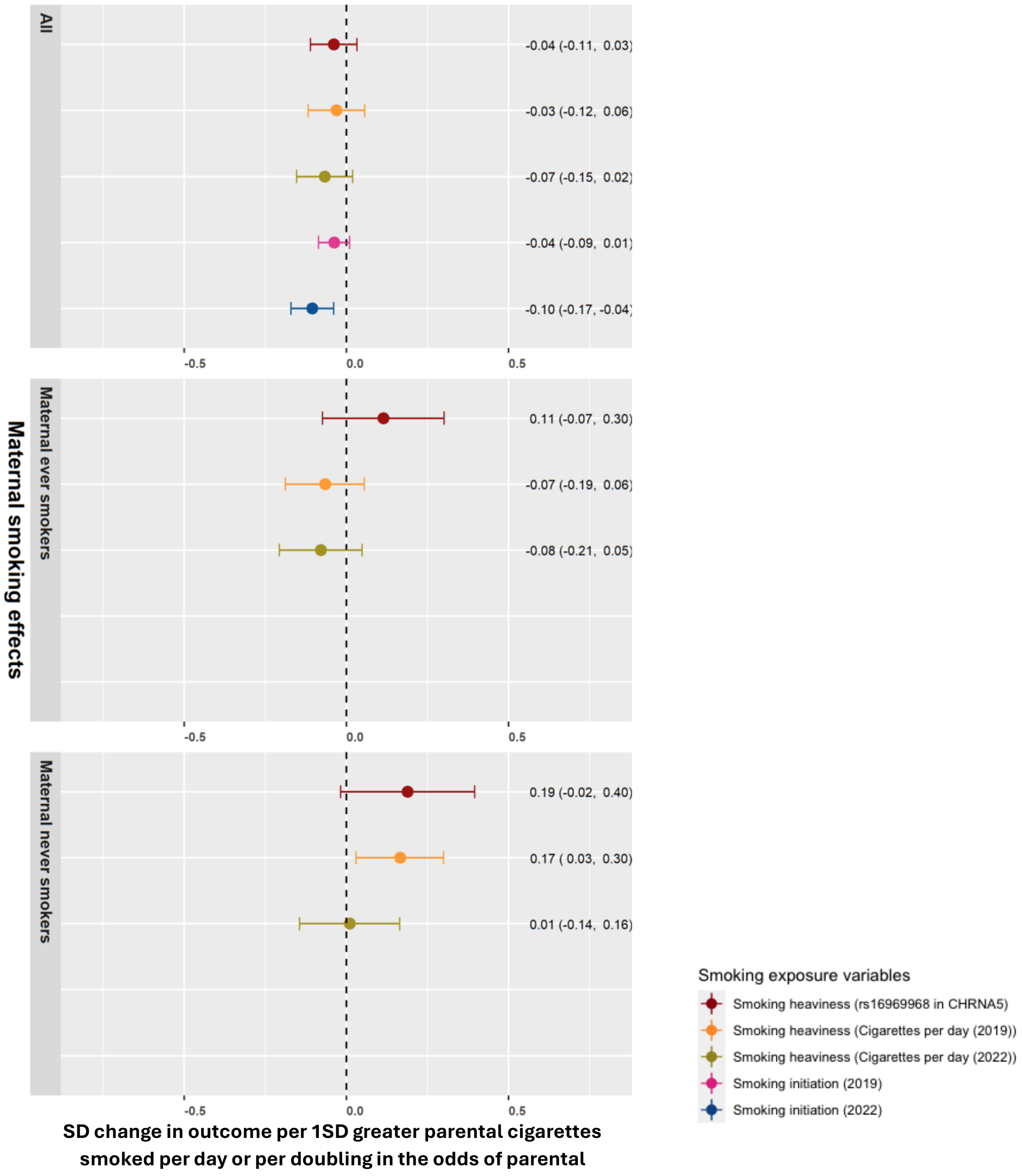
